# How to Make COVID-19 Contact Tracing Apps work: Insights From Behavioral Economics

**DOI:** 10.1101/2020.09.09.20191320

**Authors:** Ian Ayres, Alessandro Romano, Chiara Sotis

**Affiliations:** Yale Law School; Bocconi Law School; London School of Economics and Political Science

**Keywords:** COVID-19, Contact Tracing, Framing, Media

## Abstract

Due to network effects, Contact Tracing Apps (CTAs) are only effective if many people download them. However, the response to CTAs has been tepid. For example, in France less than 2 million people (roughly 3% of the population) downloaded the CTA. Against this background, we carry out an online experiment to show that CTAs can still play a key role in containing the spread of COVID-19, provided that they are re-conceptualized to account for insights from behavioral science. We start by showing that carefully devised in-app notifications are effective in inducing prudent behavior like wearing a mask or staying home. In particular, people that are notified that they are taking too much risk and could become a superspreader engage in more prudent behavior. Building on this result, we suggest that CTAs should be re-framed as Behavioral Feedback Apps (BFAs). The main function of BFAs would be providing users with information on how to minimize the risk of contracting COVID-19, like how crowded a store is likely to be. Moreover, the BFA could have a rating system that allows users to flag stores that do not respect safety norms like wearing masks. These functions can inform the behavior of app users, thus playing a key role in containing the spread of the virus even if a small percentage of people download the BFA. While effective contact tracing is impossible when only 3% of the population downloads the app, less risk taking by small portions of the population can produce large benefits. BFAs can be programmed so that users can also activate a tracing function akin to the one currently carried out by CTAs. Making contact tracing an ancillary, opt-in function might facilitate a wider acceptance of BFAs.

## 1. Introduction

Contact tracing, defined by the World Health Organization (WHO) as “the process of identifying, assessing, and managing people who have been exposed to a disease to prevent onward transmission” [1] is unanimously considered a key tool to contain the spread of COVID-19. Traditionally, contact tracing is carried out by public health authorities who interview people that have tested positive to an infectious disease. In these interviews, respondents are asked to identify the people that they have been in contact with and to determine some characteristics of these encounters (e.g. duration, proximity, etc.). After the interview, contact tracers attempt to inform the people that have been in contact with the person who tested positive and give them recommendations on how to behave.

However, the COVID-19 pandemic has proven to be a challenge for traditional contact tracing due to the speed at which the virus spreads [2], and to the fact that patients can be contagious while asymptomatic [3]. Under these circumstances, human contact tracing becomes a highly inaccurate process. On the one hand, often people cannot remember who they were in close contact with during the period in which they were infectious. On the other hand, for people it is virtually impossible to identify strangers that they have been in contact with in places like stores and public transportation. Additionally, contact tracing can only be effective if people are informed in timely manner that they have been exposed to the virus, so that they can self-isolate and avoid spreading the virus further [4]. But human contact tracing is a slow and expensive process that can take many days, if not weeks [3].

To address these shortcomings of human contact tracing, policymakers around the world have looked at Contact Tracing Apps (CTAs). In a nutshell, the basic idea is that people would download a CTA on their cellphone that can inform them when they have been in contact with a person that has tested positive for COVID-19. In principle, CTAs could be the perfect complement to human contact tracing, as they can record contacts among people that do not know each other and can send notifications almost instantaneously. Consequently, it is not surprising that almost 50 countries have either launched or are planning to launch a CTA [5].

Nevertheless, the results produced by CTAs have been disappointing in virtually all 2 places in which they have been introduced. As they are currently conceived, CTAs’ main function is to notify people when they have entered in contact with a person who tested positive to COVID-19. To perform this task, it is necessary that both the infected person and the person she entered into contact with have installed the app. Therefore, if 50% of the people download the app, only 25% of the encounters between two randomly picked members of the population would be registered. Consequently, epidemiologists have estimated that an adoption rate of 90%-95% is required in order to effectively control COVID-19 spread [6, 7]. Additionally, they have estimated that for contact tracing in general to be effective it is necessary to identify and isolate 60% of cases within a few days [4, 8].

However, even in countries characterized by high level of trust towards institutions like Singapore and Norway, governments have struggled to convince their citizens to download the CTA. In Singapore only 20% of the residents have installed the CTA [9] and in Norway the percentage of people that downloaded the app two weeks after its release was even lower (16%) [10]. Recent surveys highlight privacy, data storage and battery use concerns as the prime issues preventing people from downloading CTAs [11, 12]. Against this background, a lively debate sparked on how useful CTAs can be in preventing the spread of COVID-19. Some scholars suggest that they are an invaluable tool to contain COVID-19 diffusion [4], whereas others argue that they are largely ineffective [13, 8].

Most of these studies, however, consider people’s behavior as exogenous. That is, they implicitly assume that how often people wear a mask, how often they attend gatherings and how often they leave their home will not be affected by the existence and adoption of a CTA. In this vein, they compare the status quo with a world in which people behave in exactly the same manner, except for having downloaded a CTA. However, not anticipating behavioral responses is an important shortcoming of these studies as a large body of research shows that whether people wear a mask, attend large gatherings and practice social distancing is a fundamental determinant of how fast COVID-19 can spread [14]. In this article, we challenge this assumption and suggest that CTAs are likely to affect how people behave.

In particular, we suggest that the effect of CTAs on behaviors could go in two directions. On the one hand, a person might engage in more risky behaviors when there is a CTA that can mitigate the diffusion of the virus and inform the person when she enters in contact with someone who tested positive. This is the so-called Peltzman effect [15]. The beneficial effects of strategies aimed at reducing the level of risk to which a certain population is exposed would be offset by increased risk-taking by the target population. In other words, after installing the app, people might feel safer and hence wear masks less often or meet more people. If this hypothesis is verified, CTAs might even contribute to the diffusion of the virus.

On the other hand, in app-notifications from the CTA could reduce risk taking. In fact, insights from behavioral science suggest that carefully devised messages can induce people to engage in pro-social behavior [16, 17, 18]. The CTA could, for instance, notify the individuals when they are getting in contact with too many people, and hence might be contributing to the spread of the virus. If this hypothesis is correct, then CTAs can become more than just a complement to human contact tracing. They could become a precious communication channel between health authorities and the general public that can be used to promote prudent behaviors.

While our results offer no support to the Peltzman effect, we find that in-app notifications can be really effective tools to promote pro-social behaviors and to induce people to take more precautions. Consequently, we suggest that CTAs should be re-conceptualized as Behavioral Feedback Apps (BFAs) to account for this finding. The main function of BFAs would be providing users with information and feedback on how to behave to minimize the risk of contracting COVID-19. To carry out this function the app would *not* need to track the movements of the users but should provide users with useful information. Users, however, should be able to activate and deactivate the tracing function at any time. In this vein, contact tracing would become an ancillary service provided by BFAs, and only for users that decide to activate this function.

In particular, we suggest that BFAs should inform users on the level of risk of areas in which she is interested, based on how likely it is that the area will be crowded and on the number of cases. Some CTAs already provide similar information [19]. Additionally, the BFAs could allow users to rate businesses they visit based on whether such businesses are complying with mask-wearing and social distancing norms. Many apps that have introduced a rating system (e.g. TripAdvisors) are widely used, thus suggesting that people are inclined to engage with rating systems. From this perspective, the app would serve five purposes. First, it would directly protect the app user by giving information on which areas and which businesses to avoid. Second, it would incentivize business owners and their employees to respect social distancing rules to avoid negative reviews. Third, it would facilitate government enforcement, since public authorities would be warned about businesses that are recidivist violators of norms aimed at minimizing the spread of COVID-19. Fourth, the app would protect the community because people following its advice will reduce the potential for contacts in crowded areas. Last, using the app in this way could help creating a bond between the users and the app, which might increase the likelihood that the user will activate the tracing function.

The tracing function would work like present-day CTAs, but would also include tailored in-app notifications like the ones in Fig. 3. Consequently, the app could send information as “You are taking less risks than your peers. You are protecting your community” or “You are among the users that took the most risks. You could become a superspreader”.

Importantly, BFAs would provide useful guidance and feedback even if the percentage of people that download it and activate the tracing function is relatively small. In fact, behavioral guidance does not require a critical mass of downloads comparable to that required by contact tracing.

## 2. Background Information

### 2.1. Contact Tracing Apps Around the World

Singapore was the first country to introduce a CTA. In March 2020 it released TraceTogether, which uses Bluetooth technology to inform users when they have been in contact with someone who tested positive for COVID-19 [20]. The results produced by TraceTogether have not been fully satisfying. Only 20% of the residents downloaded it [9], and the app has some problems with Apple operating systems (iOS) [20]. Despite these issues, many countries followed Singapore’s lead and implemented their own app. For instance, Australia introduced the app CovidSafe, which shares many features with TraceTogether. To this date, over 6 million people – or about one quarter of the population – have downloaded it. But many have questioned its utility, given that the numbers of cases that it helped identifying appears to be small. One of the reasons for this is that the app does not register proximity between two cellphones if the screen is locked [12].

Also many European countries have embraced enthusiastically the idea of using CTAs to complement the efforts of their healthcare workers. In June, France, Germany and Italy, the three biggest economies of continental Europe, all launched their version of a CTA.

France released the app StopCovid in June, but two months after its release less than two million people – or roughly 3% of the population – had downloaded it [21]. In an attempt to increase the number of users, French authorities attempted to inform the public about the importance of the app. For instance, Olivier Ve’ran, France Health Ministry, tweeted that people can use StopCovid to protect themselves and the people they meet during their holidays. Meanwhile, however, hundreds of thousands of people seem to have uninstalled the app because it consumes too much battery [12].

In Germany the introduction of a CTA was sponsored from the very beginning by the most important political leaders. In an official video-message, Angela Merkel said that the Corona-Warn app is a companion and protector and an important helper that will benefit the community [22]. The spokesman of the German government also stressed that downloading the app is both in the interest of the individuals and of the community and that the app can be an effective tool to help containing the spread of COVID-19 [23]. In response to these calls roughly 16 million people have downloaded Corona-Warn, which is about 20% of the population.

In a similar fashion, key figures of the Italian government spoke in support of the Italian CTA Immuni. The Italian prime minister Giuseppe Conte, in an official message to the citizens, noted that the app is safe and that it respects the user’s privacy [24]. Like his European counterparts he emphasized that downloading the app is entirely voluntary, but that by downloading the app citizens can help containing the spread of the virus [25]. By the 5th of August 2020, Immuni had been downloaded 4.6 million times. Moreover, according to the Italian Ministry of Innovation it had helped breaking the chain of contagions in two key circumstances by allowing health authorities to identify 63 people that were positive to COVID-19 [26].

In the United States, the Federal government did not show the same kind of support to CTAs. There have been a number of attempts but, even when they were promoted by tech giants like Google, CTAs failed to reach a critical mass of users [27]. At the state level, however, there is more support for these apps. For instance, the Governor of Virginia has strongly endorsed COVIDWISE, which is the first statewide app to use the technology developed by Google and Apple. He argued that the app “can really help us catch new cases early before they spread as far” [28]. Alabama, North Dakota and South Carolina also expressed interest in the technology developed by Google and Apple as a means to contain the spread of the virus [29]. Rhode Island went in a different direction, instead, and supported the use of the “homegrown app” Crush COVID RI, which, according to Governor Raimondo, is a “a tool that helps everybody in Rhode Island get through the crisis” [30]. The Governor also emphasized that the app “protects people’s privacy and data in an ironclad way” [30]. About 60.000 people decided to download the app (about 5% of the population), while Raimondo is hoping to reach 100.000 downloads. Other States like North Dakota, South Dakota, Utah and Wyoming also developed their version of the app.

However, citizens’ response has been tepid at best: in North Dakota and in Utah respectively less than 5% and 3% of the population downloaded the app [31], a far cry from the critical mass epidemiologists suggest is needed to make it useful. The result was so disappointing that Utah authorities decided to turn off the location tracking function, and to use the app only as a way to communicate COVID-19 related information with the population.

This brief overview reveals that policymakers are keenly aware of the importance of reaching a critical mass of downloads, but reaching that critical mass has proven to be problematic. For this reason, most of the communication from political leaders and health authorities has been aimed at increasing the number of downloads by emphasizing the role that the CTA can play in containing the spread of the virus and by reassuring the public that CTAs respect users’ privacy.

### 2.2. Literature Review

The literature on CTAs has mushroomed during the COVID-19 pandemic. Studies have focused around three main issues. First, many investigated how to best develop and improve these apps from a technical perspective e.g. [32]. A second group of studies analyzed the privacy problems posed by CTAs [33], and how to minimize them [34]. A last strand of literature investigated what influences people decision to download the app. For instance, Abeler et al [35] find that people might resist downloading the app because: (i) they fear that the government might keep monitoring citizens also after the pandemic is over, (ii) they are concerned that it might make them more anxious and (iii) they are afraid that their cellphone might get hacked. Frimpong and Helleringer [36] study how financial incentives influence the decision to download the app and find that they can be effective. It is unclear, however, how to ensure that people do not uninstall the CTA after receiving the financial incentive. Kaptchuck et al. [37], instead, carry out a survey with 4500 Americans to investigate which attributes of the app they value the most. They find that ensuring that the app is “perfectly private” greatly increases the number of people who are willing to install it. They also find that Americans value how much the app can contribute to public health, and that false negatives have a larger effect than false positives on reducing the willingness to download the app. Last, using a sample of 1117 Australians, Bradshaw et al. [11] find that data safety is an important determinant of whether people download the app.

Notwithstanding the importance of these works, here we focus on a different issue that to the best of our knowledge has been largely overlooked. Specifically, we study whether people’s behavior is affected by the information governments give about the app in the pre-download phase, and by the in-app notifications in the post-download phase.

Thinking about the communication by public authorities in the pre-download phase, it is possible that excessively positive descriptions of the app could induce too much optimism in the general public. Consequently, people might feel protected by the app and engage in activities that increase the risk of contracting the virus like attending small and large gatherings. This would be in line with the risk compensation theory, which posits that when people feel more protected they will engage in more risky behaviors. For example, [38] and [39] find that the presence of pre-exposure prophylaxis – a strategy that helps protecting against HIV – induces people to engage in more condomless sex. There is some evidence in support of the risk compensation theory also in the context of COVID-19. Cartaud et al. [40] find that people wearing mask preferred a lower interpersonal distance than people without masks. Similarly, Yan et al. [41] find that people living in American States that mandated the use of masks spent less time at home and visited more commercial businesses. If the risk compensation theory holds true in this context, it would pose a dilemma for health authorities: promoting the app emphasizing only its advantages, like the German government did, may induce more people to download it. However, people might then engage in more risky activities. Vice versa, if health authorities flag also the downsides of the app, people might refrain from engaging in more risky activities, but the number of downloads might be too small for the app to become an effective tool in containing the spread of the disease.

Additionally, the app opens a communication channel between the health authorities and the app users. Health authorities can then use in-app messages to induce people to engage in pro-social behavior. For example, Ayres et al. [42] and Allcott [43] find that informing people when they are consuming more energy than their neighbors can lead to considerable savings. Additionally, Ferraro and Price find that a similar nudge can help reducing water consumption [44]. Therefore, we hypothesize that informing people about how much risk they are taking in comparison to other users might induce app users to engage in more pro-social behaviors like wearing mask or staying home. In the context of COVID-19, researchers are converging on the idea that 20% of the people cause 80% of all COVID-19 cases (i.e. superspreaders) [45, 46]. Consequently, we hypothesize that informing people that they might be a superspreader could be an effective way to induce them to engage in more pro-social behavior.

## 3. Materials and Methods

We devised a double-blind experiment approved by the Yale IRB divided in two phases. Participants were informed at the beginning of Phase I that the experiment was composed of two phases. Only those who participated in Phase I would be allowed to take part in Phase II. Participants were paid $1 for taking part in Phase I and $0.75 for taking part in Phase II. In addition, we also informed them that by participating in both Phases they would be automatically entered into a lottery with three prizes of $100 and one of $200.

Phase I aimed at studying the impact on behaviors of various kinds of communication strategies in the pre-download phase. Consequently, the underlying assumption is that participants do not have the CTA on their cellphone. Participants in the treatment groups are then provided with information about the app, devised to mimic governments’ campaigns that were run around the world. Phase II was carried out one week after Phase I and aimed at studying the impact of in-app notifications on behaviors. Therefore, participants were asked to imagine that they had downloaded the app, and were exposed to different kinds of in-app notifications. To improve the degree of realism of the experiment, all the in-app notifications use a similar graphic to the Italian CTA Immuni. We built on this CTA because its developers have shared all their code and screenshots of the notifications on GitHub.

### 3.1. Phase I: Design

We recruited a sample of *n* = 1500 U.S. residents on Prolific. In Phase I respondents were randomly assigned to one of three different groups: Pros, Pros and Cons and Control I. The respondents included in the Pros group were shown information on the advantages of CTAs (Fig. 1). The respondents in the Pros and Cons group were shown information on both the advantages (Fig. 1) and the problems that characterize CTAs (Fig. 2). Last, respondents in Control I group were not given any information on CTAs. We then asked the same questions to all participants.

**Figure 1:**
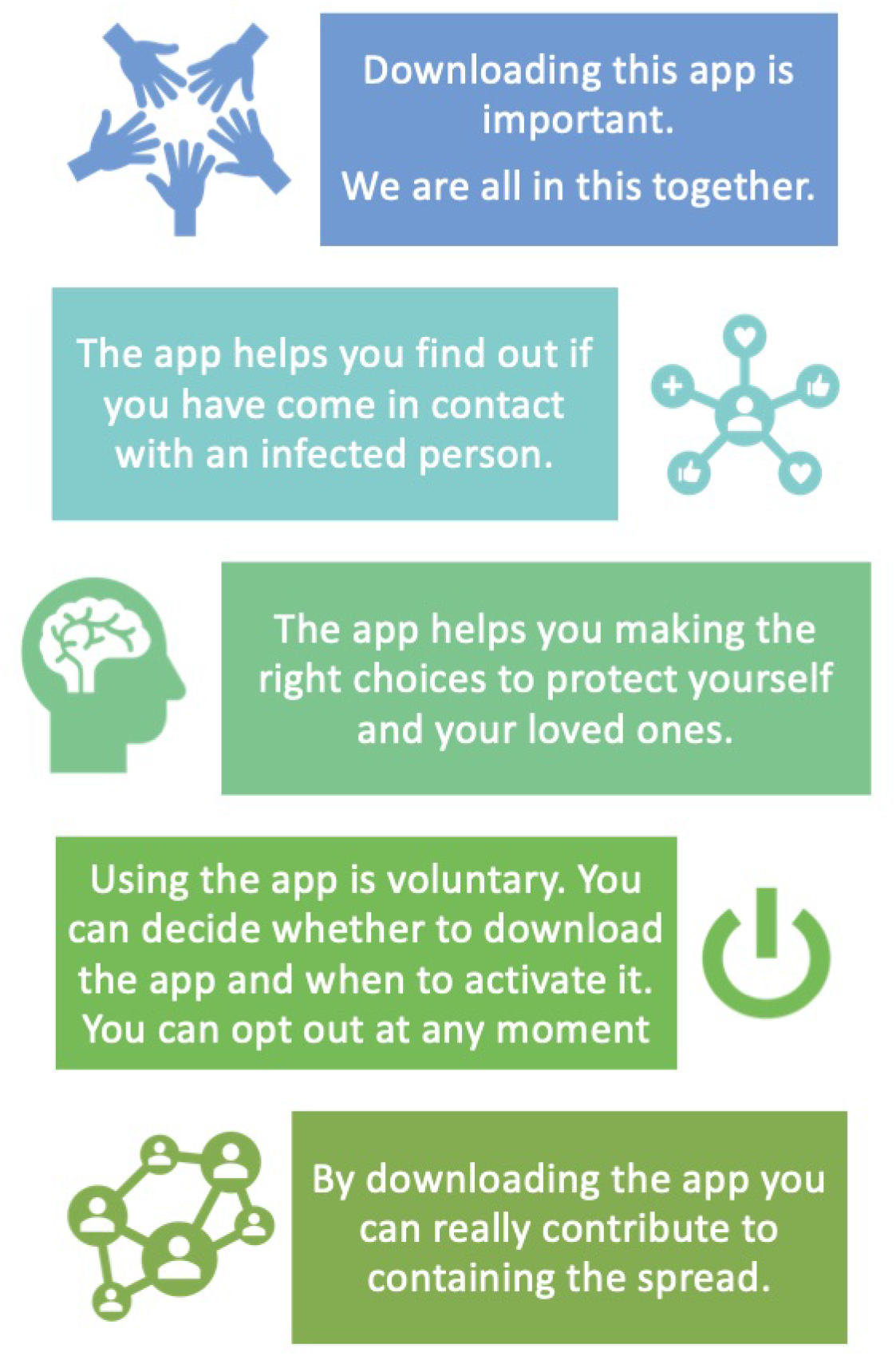
Treatment I, Phase I: Participants see some of the app’s pros.

**Figure 2:**
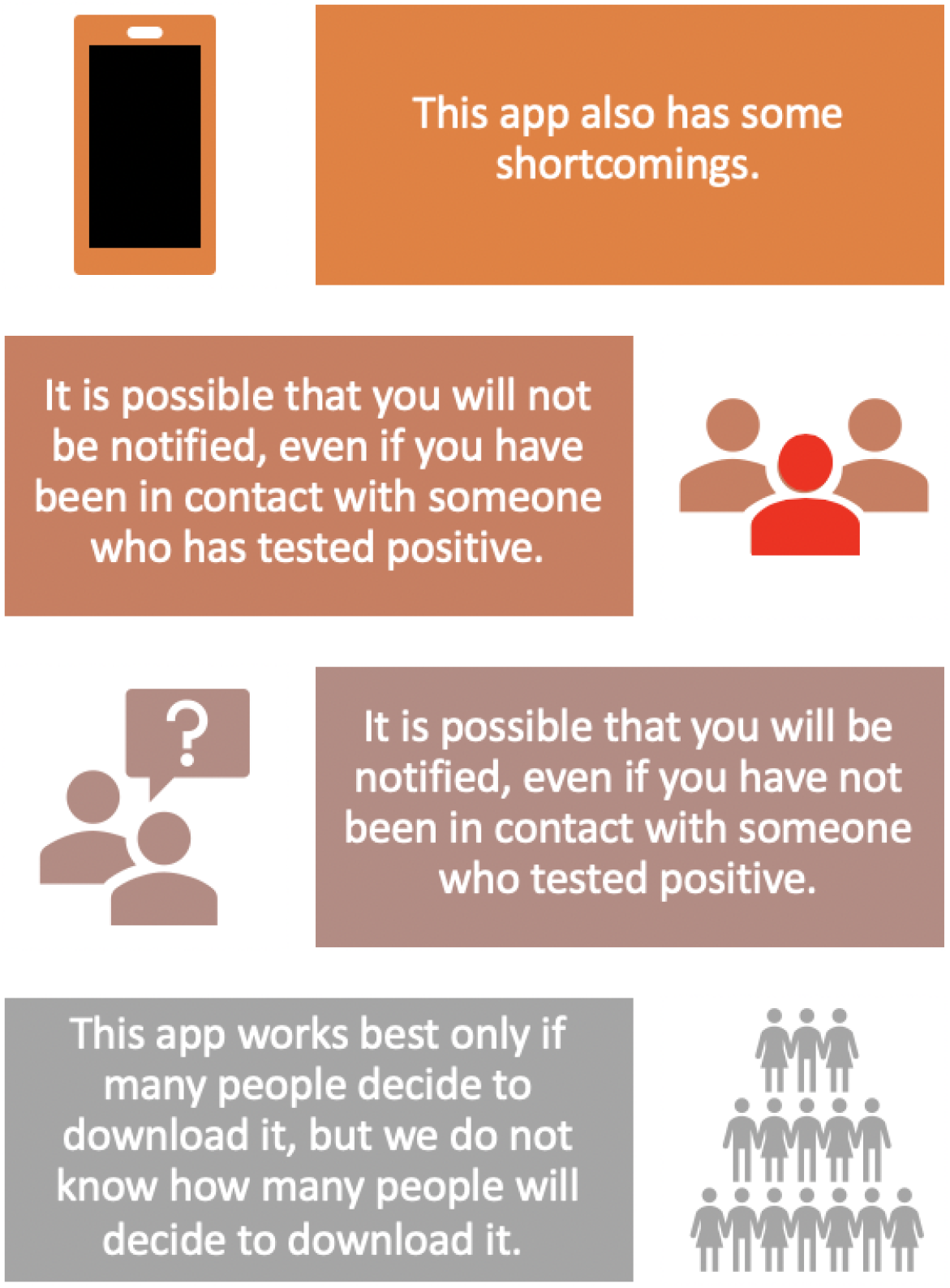
Treatment II, Phase I: Participants see some of the pros and cons of the app.

To begin with, we asked respondents how likely they were to engage in some behaviors (outside of the workplace) in the coming week (hereinafter “behavioral questions”). In particular, we asked how likely they were to: (*i*) attend small gatherings, (*ii*) attend large gatherings, (*iii*) see older relatives or people at risk, (*iv*) stay home unless they need to go for groceries/shopping for essential items, and (*v*) wear a mask. In the following screen we asked them how worried they are of contracting COVID-19. All these questions were asked on a Likert scale from 1 to 10 where 1 meant “Absolutely certain that you will not engage in this activity” and 10 “Absolutely certain that you will engage in this activity”. After this, we asked respondents how likely they were to download the app (5-point Likert scale from “Extremely Unlikely” to “Extremely Likely”). To the people that said that they were neither likely nor unlikely, unlikely, or extremely unlikely to download the app we also asked the amount of money which would make them willing to accept downloading the app. We then asked which percentage of people they thought were going to download the app, and whether they agreed that CTAs were important to contain the spread of COVID-19 (5-point Likert scale from “Strongly Disagree” to “Strongly Agree”).

Lastly, we asked standard demographic questions and other questions related with how much respondents trust various actors, and whether they thought that their State and the Federal Government had handled the COVID-19 crisis properly. We asked trust towards: people known personally, people not known personally, US government, health insurance companies, the media, the CDC and the healthcare system. As these measures are all related and can affect the response to the treatments, we used principal component analysis to construct a trust index.

### Phase II: Design

A week later we carried out Phase II. Out of the initial 1500 participants from the first round 1303 also completed Phase II. In this part of the experiment we first asked participants how they behaved the previous week using the same behavioral questions asked in Phase I. Then we asked participants to imagine that they had downloaded the app. Participants were then randomly assigned to four groups: Good Behavior, Bad Behavior, Superspreader and Control II. Fig. 3 shows the three treatments.

**Figure 3:**
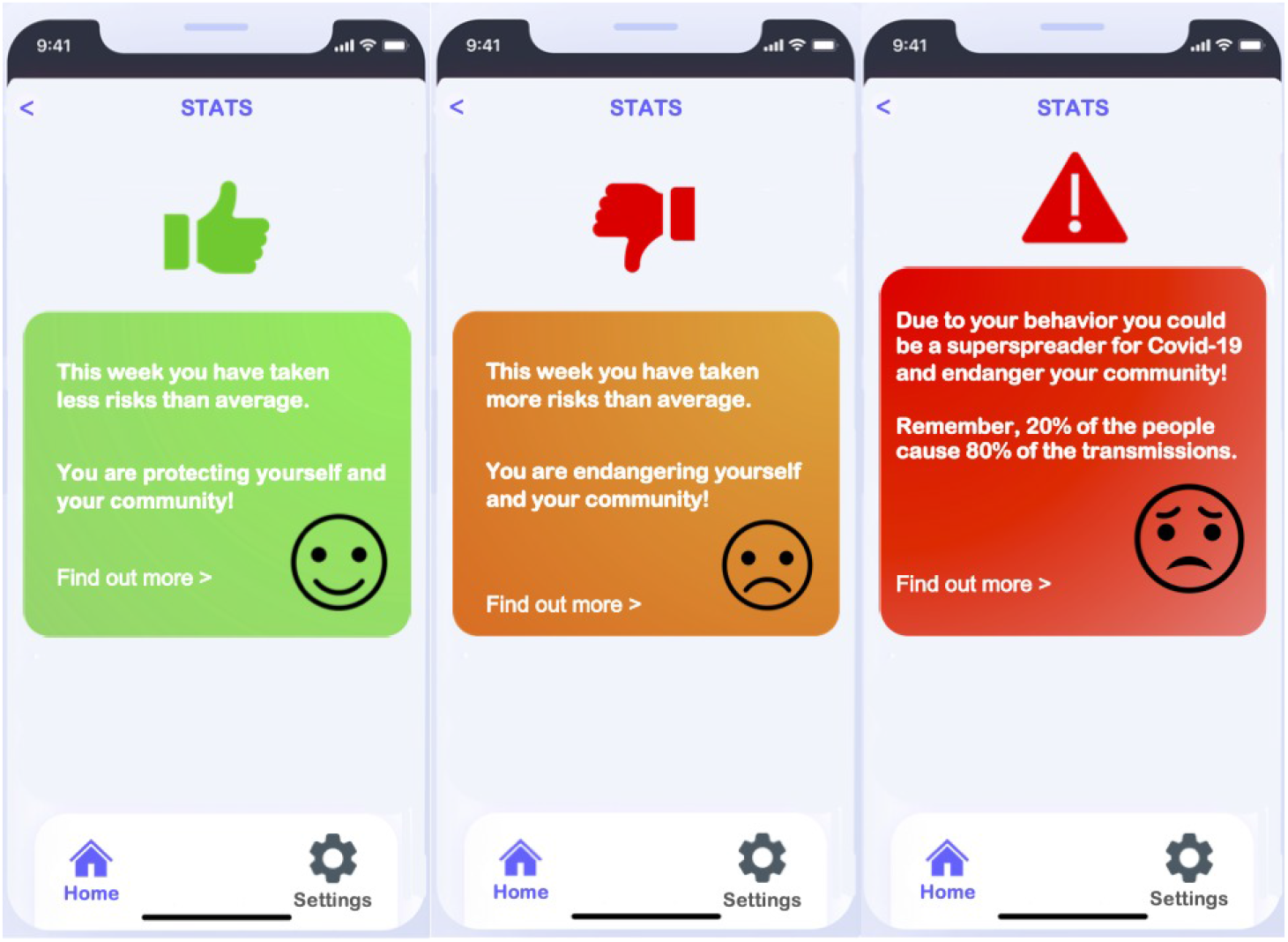
Treatment I, Phase II: Participants are informed that they are taking less risk than the average user and protecting their community (Left Panel). Treatment II, Phase II: Participants are informed that they are taking more risk than the average person and endangering their community (center panel). Treatment III, Phase II: Participants are informed that due to their behavior they might become superspreaders (right panel)

The participants in the Good Behavior group were shown an in-app notification that read “This week you have taken less risks than average. You are protecting yourself and your community” on a green screen with a smiling face. The participants in the Bad Behavior group were shown an in-app notification that read “This week you have taken more risks than average. You are endangering yourself and your community” on an orange screen with an unhappy face. The participants in the Superspreader group received a notification that read “Due to your behavior you could be a superspreader for Covid-19 and endanger your community. Remember 20% of the people cause 80% of transmissions” on a red screen with a worried face. Last, the respondents in Control II group were not shown any notification. After this screen, we asked to all participants how they intended to behave in the coming week using the same behavioral questions asked in Round I. We also asked them how long for they were planning to keep the app.

Afterwards, we showed to all participants a notification that they had exposed to the virus (Fig. 4 left panel), and another screen suggesting how to behave following the contact (Fig. 4 right panel). In particular, the app suggested to: (*i*) wash hands frequently with soap, (*ii*) cough and sneeze into a a tissue or in the crook of the elbow, (*iii*) assess symptoms and take temperature twice per day and (*iv*) stay home and practice social distancing for 14 days since the contact. These recommendations were all based on those given by the Italian app Immuni and are in line with suggestions from the WHO and US Center for Diseases Control and Prevention (CDC) [47]. We then asked respondents on a Likert scale from 1 to 10 how likely they were to follow each recommendation of the app.

**Figure 4:**
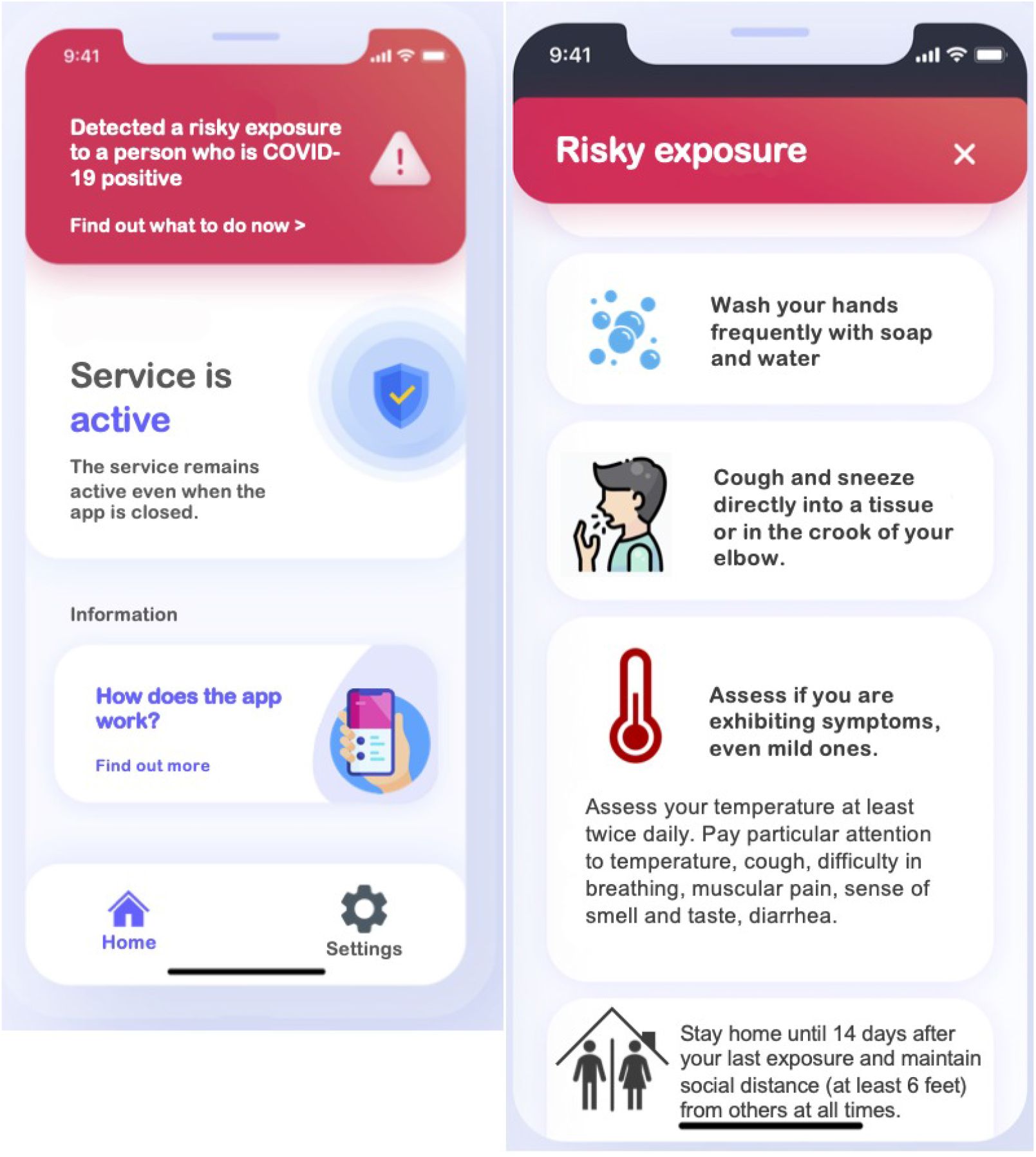
Warning message for a potential contact (left panel). The recommendations given by the app after the warning (right panel)

Last, we turned to questions related to the prevention optimism based on the CTA and asked respondents how much they agreed with certain statements (see Table 1). These statements were adapted from the medical literature on prevention optimism for HIV related to Pre-exposure Prophylaxis [38].

**Table 1:** The table summarizes the effects of the various treatments on the level of agreements with statements aimed at identifying prevention optimism. * significant at 10%, ** significant at 5% and *** significant at 1%.

| Statement | Agrees More | Agrees Less |
| --- | --- | --- |
| Because of the contact tracing app I am less likely to contract COVID-19 | Good Behavior** | Pros and Cons** |
| It is safe for me to be around people because of the contact tracing app | - | Pros and Cons**,<br>Bad Behavior**,<br>Superspreaders** |
| Because of the contact tracing app less people will be infected by COVID-19 | Good Behavior** | - |
| If many people download the contact tracing app, wearing a mask becomes less important | - | Pros and Cons* |

## 4. Results of Phase 1

In Phase 1 respondents are randomly assigned to three groups: Pros, Pros and Cons and Control I. To the Pros group we showed the pros of the app (Fig. 1), to the Pros and Cons we showed both the strengths and the weaknesses of the app (Fig. 2), while we provided no information on the app to Control I group.

We observe that participants in the treated groups are less worried about the pandemic (Table 5). The result is robust and significant at 1% for the Pro group, whereas it is smaller, less robust and significant only at 10% for the Pros and Cons group. These results are consistent with the idea that a CTA can be reassuring. In fact, the effect is stronger if only the advantages of the app are emphasized.

We observe that the two treatments have no significant effects on behaviors. This suggests that communicating just the pros of the app, or also its cons, will not influence how people behave after having downloaded the app.

**Table 5:** Determinants of worry: combined controls, ordered logit estimates, robust standard errors

|  | (1) | (2) | (3) | (4) |
| --- | --- | --- | --- | --- |
|  | Worry About Covid-19 | Worry About Covid-19 | Worry About Covid-19 | Worry About Covid-19 |
| Worry About Covid-19 |  |  |  |  |
| Treatment 1: Only Positive Info | -0.418***<br>(0.001) | -0.378***<br>(0.004) | -0.418***<br>(0.001) | -0.378***<br>(0.004) |
| Treatment 2: Positive and Negative Info | -0.230*<br>(0.083) | -0.188<br>(0.167) | -0.230*<br>(0.083) | -0.188<br>(0.167) |
| essential | 0.577***<br>(0.000) | 0.584***<br>(0.000) | 0.577***<br>(0.000) | 0.584***<br>(0.000) |
| econsituation | -0.348***<br>(0.000) | -0.330***<br>(0.000) | -0.348***<br>(0.000) | -0.330***<br>(0.000) |
| Male | -0.184<br>(0.102) | -0.179<br>(0.125) | -0.184<br>(0.102) | -0.179<br>(0.125) |
| Education | 0.0512<br>(0.252) | 0.0332<br>(0.488) | 0.0512<br>(0.252) | 0.0332<br>(0.488) |
| Age | 0.0120***<br>(0.008) | 0.0128***<br>(0.007) | 0.0120***<br>(0.008) | 0.0128***<br>(0.007) |
| Income | -0.0134<br>(0.435) | -0.0222<br>(0.229) | -0.0134<br>(0.435) | -0.0222<br>(0.229) |
| stateresponse | -0.0759*<br>(0.093) | -0.103**<br>(0.030) | -0.0759*<br>(0.093) | -0.103**<br>(0.030) |
| USresponse | -0.191***<br>(0.002) | -0.168***<br>(0.007) | -0.191***<br>(0.002) | -0.168***<br>(0.007) |
| likelydownload | 0.261***<br>(0.000) | 0.266***<br>(0.000) | 0.261***<br>(0.000) | 0.266***<br>(0.000) |
| importance | 0.249***<br>(0.000) | 0.225***<br>(0.001) | 0.249***<br>(0.000) | 0.225***<br>(0.001) |
| Usdownloads | 0.0135***<br>(0.000) | 0.0104***<br>(0.002) | 0.0135***<br>(0.000) | 0.0104***<br>(0.002) |
| trustUSgov | -0.0160<br>(0.646) | -0.0383<br>(0.296) | -0.0160<br>(0.646) | -0.0383<br>(0.296) |
| trustinsurance | 0.0362<br>(0.297) | 0.0113<br>(0.765) | 0.0362<br>(0.297) | 0.0113<br>(0.765) |
| trustCDC | 0.0837***<br>(0.003) | 0.0495<br>(0.120) | 0.0837***<br>(0.003) | 0.0495<br>(0.120) |
| Republican |  | 0.307*<br>(0.081) |  | 0.307*<br>(0.081) |
| Democrat |  | 0.270**<br>(0.033) |  | 0.270**<br>(0.033) |
| householdsize |  | 0.0714<br>(0.133) |  | 0.0714<br>(0.133) |
| trustmedia |  | 0.0822***<br>(0.007) |  | 0.0822***<br>(0.007) |
| trusthealthcare |  | 0.0169<br>(0.579) |  | 0.0169<br>(0.579) |
| / |  |  |  |  |
|  | (0.000) | (0.000) | (0.000) | (0.000) |
| Observations | 1158 | 1126 | 1158 | 1126 |

p-values in parentheses
\* $p < 0.10$ , \*\* $p < 0.05$ , \*\*\* $p < 0.01$

At the same time, we find that both treatments increase the likelihood to download the app (Table 6). Notably, the result is more robust for the Pros and Cons treatment, suggesting that communicating also the downsides of the app can promote downloads. This suggests that governments that only emphasized the virtues of CTAs might have chosen the wrong communicative strategy if their goal was to maximize the number of downloads.

**Table 6:** Determinants of download: combined controls, ordered logit estimates, robust standard errors

|  | (1) | (2) | (3) | (4) | (5) | (6) |
| --- | --- | --- | --- | --- | --- | --- |
|  | likelydownload | likelydownload | likelydownload | likelydownload | likelydownload | likelydownload |
| likelydownload |  |  |  |  |  |  |
| Treatment 1: Only Positive Info | 0.247* | 0.217 | 0.213 | 0.242* | 0.185 | 0.178 |
|  | (0.076) | (0.124) | (0.133) | (0.085) | (0.201) | (0.220) |
| Treatment 2: Positive and Negative Info | 0.288** | 0.260* | 0.277* | 0.312** | 0.256* | 0.265* |
|  | (0.038) | (0.067) | (0.052) | (0.026) | (0.078) | (0.068) |
| Worry About Covid-19 | 0.140*** | 0.136*** | 0.123*** | 0.135*** | 0.130*** | 0.120*** |
|  | (0.000) | (0.000) | (0.000) | (0.000) | (0.000) | (0.000) |
| essential | -0.193 | -0.190 | -0.209 | -0.175 | -0.168 | -0.183 |
|  | (0.179) | (0.194) | (0.164) | (0.222) | (0.260) | (0.232) |
| econsituation | -0.0662 | -0.0688 | -0.0761 | -0.0959 | -0.0932 | -0.0946 |
|  | (0.440) | (0.425) | (0.379) | (0.267) | (0.289) | (0.286) |
| Male | 0.113 | 0.152 | 0.123 | 0.0391 | 0.0573 | 0.0449 |
|  | (0.340) | (0.208) | (0.321) | (0.750) | (0.652) | (0.727) |
| Education | 0.0529 | 0.0571 | 0.0485 | 0.0352 | 0.0354 | 0.0314 |
|  | (0.228) | (0.205) | (0.290) | (0.439) | (0.457) | (0.518) |
| Age | -0.0155*** | -0.0146*** | -0.0141*** | -0.0154*** | -0.0148*** | -0.0143** |
|  | (0.002) | (0.006) | (0.010) | (0.003) | (0.007) | (0.011) |
| Income | 0.0355* | 0.0284 | 0.0217 | 0.0322* | 0.0257 | 0.0206 |
|  | (0.060) | (0.154) | (0.286) | (0.096) | (0.213) | (0.328) |
| stateresponse | 0.0157 | 0.00816 | 0.00310 | 0.00774 | -0.0182 | -0.0228 |
|  | (0.749) | (0.870) | (0.951) | (0.876) | (0.723) | (0.661) |
| USresponse | 0.0498 | 0.0701 | 0.0720 | -0.0654 | -0.0653 | -0.0552 |
|  | (0.387) | (0.276) | (0.263) | (0.399) | (0.420) | (0.492) |
| importance | 1.114*** | 1.111*** | 1.093*** | 1.074*** | 1.096*** | 1.087*** |
|  | (0.000) | (0.000) | (0.000) | (0.000) | (0.000) | (0.000) |
| Usdownloads | 0.0299*** | 0.0303*** | 0.0294*** | 0.0289*** | 0.0293*** | 0.0285*** |
|  | (0.000) | (0.000) | (0.000) | (0.000) | (0.000) | (0.000) |
| Republican |  | -0.00770 | -0.0550 |  | -0.0339 | -0.0784 |
|  |  | (0.967) | (0.772) |  | (0.861) | (0.694) |
| Democrat |  | 0.201 | 0.193 |  | 0.0983 | 0.0950 |
|  |  | (0.132) | (0.149) |  | (0.477) | (0.492) |
| householdsize |  | 0.0510 | 0.0401 |  | 0.0351 | 0.0244 |
|  |  | (0.261) | (0.387) |  | (0.441) | (0.601) |
| smallmeet |  |  | 0.00635 |  |  | 0.00324 |
|  |  |  | (0.806) |  |  | (0.903) |
| largemeet |  |  | 0.0418 |  |  | 0.0367 |
|  |  |  | (0.274) |  |  | (0.346) |
| seeatrisk |  |  | 0.0302 |  |  | 0.0283 |
|  |  |  | (0.227) |  |  | (0.271) |
| Likelihood to Wear Masks Next Week |  |  | 0.0753* |  |  | 0.0640 |
|  |  |  | (0.058) |  |  | (0.123) |
| stayhome |  |  | 0.00795 |  |  | 0.00208 |
|  |  |  | (0.761) |  |  | (0.939) |
| trustUSgov |  |  |  | 0.0503 | 0.0610 | 0.0562 |
|  |  |  |  | (0.206) | (0.138) | (0.182) |
| trustinsurance |  |  |  | 0.0565 | 0.0412 | 0.0390 |
|  |  |  |  | (0.106) | (0.292) | (0.324) |
| trustCDC |  |  |  | 0.0493* | 0.0289 | 0.0273 |
|  |  |  |  | (0.088) | (0.376) | (0.415) |
| trustmedia |  |  |  |  | 0.0209 | 0.0171 |
|  |  |  |  |  | (0.524) | (0.605) |
| trusthealthcare |  |  |  |  | 0.0321 | 0.0312 |
|  |  |  |  |  | (0.304) | (0.325) |
| / |  |  |  |  |  |  |
| Observations | 1169 | 1150 | 1144 | 1158 | 1126 | 1121 |
| p-values in parentheses |  |  |  |  |  |  |
| * $p < 0.10$, ** $p < 0.05$, *** $p < 0.01$ | | | | | | |

In line with previous papers analyzing the drivers of adoption [48, 49, 33], we find that people with higher levels of trust are more willing to download the app. We also find that treatments have different effects depending on the political ideology of the respondent. In particular, we find that republicans in the treatment groups are less willing download the app.

Notably, we find that about 56% of our respondents are willing to download the app even without a payment. Moreover, we observe that respondents stating that they are less likely to download the app want more money to download the app.

## 5. Phase 2

In Phase 2 the participants were randomly assigned to three treatment groups (Fig. 3 Good Behavior, Bad Behavior, Superspreader and Control II group.

### 5.1. Behaviors after Phase I

Respondents are first asked about their behaviors in the previous week, with the same behavioral questions asked the week before. In addition we asked two more questions on how many contacts they had during the last 7 days. We do not find differences in behavior among groups, with the exception of masks use. People who were in the treatment groups wore masks more often than the control group (significant at 5%, or 1% depending on the sets of controls) (Table 7). Overall, the main finding is that communication in the pre-download phase affects how worried people are, but has a small impact on how they behave.

**Table 7:** Determinants of wearing a mask: combined controls, robust standard errors

|  | (1) | (2) | (3) | (4) | (5) |
| --- | --- | --- | --- | --- | --- |
|  | LWmask | LWmask | LWmask | LWmask | LWmask |
| LWmask |  |  |  |  |  |
| prosonly | 0.446***<br>(0.009) | 0.465***<br>(0.007) | 0.421**<br>(0.014) | 0.443**<br>(0.011) | 0.153<br>(0.468) |
| prosandcons | 0.370**<br>(0.031) | 0.347**<br>(0.044) | 0.387**<br>(0.025) | 0.357**<br>(0.039) | 0.386*<br>(0.077) |
| worry | 0.207***<br>(0.000) | 0.202***<br>(0.000) | 0.196***<br>(0.000) | 0.194***<br>(0.000) | 0.124***<br>(0.001) |
| essential | -0.362**<br>(0.022) | -0.338**<br>(0.035) | -0.351**<br>(0.027) | -0.325**<br>(0.044) | -0.231<br>(0.268) |
| econsituation | 0.0375<br>(0.694) | 0.0225<br>(0.818) | 0.0609<br>(0.526) | 0.0547<br>(0.581) | 0.0404<br>(0.739) |
| male | -0.304**<br>(0.033) | -0.251*<br>(0.086) | -0.289**<br>(0.047) | -0.186<br>(0.220) | -0.188<br>(0.314) |
| edu | -0.00442<br>(0.934) | -0.0161<br>(0.772) | 0.00706<br>(0.896) | -0.00250<br>(0.965) | -0.00235<br>(0.973) |
| age | 0.00668<br>(0.233) | 0.00613<br>(0.295) | 0.00770<br>(0.184) | 0.00660<br>(0.279) | 0.00254<br>(0.751) |
| income | -0.00221<br>(0.918) | -0.000787<br>(0.973) | 0.0000929<br>(0.997) | -0.00350<br>(0.882) | -0.0261<br>(0.356) |
| stateresponse | -0.0293<br>(0.603) | -0.0309<br>(0.589) | -0.0555<br>(0.334) | -0.0515<br>(0.377) | -0.0443<br>(0.539) |
| USresponse | -0.189***<br>(0.002) | -0.172**<br>(0.017) | -0.0149<br>(0.856) | -0.0269<br>(0.759) | -0.0182<br>(0.871) |
| likelydownload | 0.0391<br>(0.609) | 0.0242<br>(0.755) | 0.0597<br>(0.452) | 0.0460<br>(0.574) | 0.0569<br>(0.565) |
| importance | 0.162**<br>(0.027) | 0.165**<br>(0.025) | 0.134*<br>(0.072) | 0.148*<br>(0.052) | 0.239**<br>(0.022) |
| Usdownloads | -0.00325<br>(0.430) | -0.00362<br>(0.392) | -0.00220<br>(0.606) | -0.00220<br>(0.619) | -0.00100<br>(0.855) |
| republican |  | 0.317<br>(0.166) |  | 0.441*<br>(0.064) | 0.370<br>(0.223) |
| democrat |  | 0.465***<br>(0.003) |  | 0.487***<br>(0.003) | 0.481**<br>(0.016) |
| householdsize |  | -0.0175<br>(0.762) |  | -0.0163<br>(0.776) | 0.0220<br>(0.753) |
| trustUSgov |  |  | -0.0893**<br>(0.040) | -0.0966**<br>(0.036) | -0.0841<br>(0.140) |
| trustinsurance |  |  | -0.0377<br>(0.365) | -0.0387<br>(0.384) | 0.0420<br>(0.422) |

### 5.2. In-app Notifications

Behavioral feedback messages have a significant effect on how people behave. Here we have three treatments and one control group. To one group we show a green screen saying that they have taken less risks than average (Good Behavior), to one group we show an orange screen saying that they have taken more risks than average (Bad Behavior) and to the last group we show a red screen saying that due to their behavior they might be super-spreaders (Superspreaders) and explain what super-spreaders are (Fig. 3).

We find that:

- *Large Gatherings*: Respondents in the Superspreaders group attend less gatherings than any other group. The result is significant at 5% (Table 8). There are no significant differences between Respondents in Good and Bad Behavior and Control group.
- *Small Gatherings*: Respondents in the Superspreaders group attend less small gatherings than any other group (significant at 1 %) (Table 9). No significant impact of the Good and the Bad behavior treatments.
- *Seeing people at risk*: Respondents in the Superspreaders group see less people at risk than any other group (significant at 1 %) (Table 10). No significant impact of the Good and the Bad behavior treatments.
- *Stay Home*: Respondents in the Superspreaders group stay at home more often than any other group (significant at 1 %) (Table 11). No significant impact of Good and the Bad behavior treatments.
- *Masks*: The Superspreaders treatment has a positive effect on mask wearing, but the statistical significance depends on the set of controls used (ranging from 1 % to not significant) (Table 12).

**Table 8:**
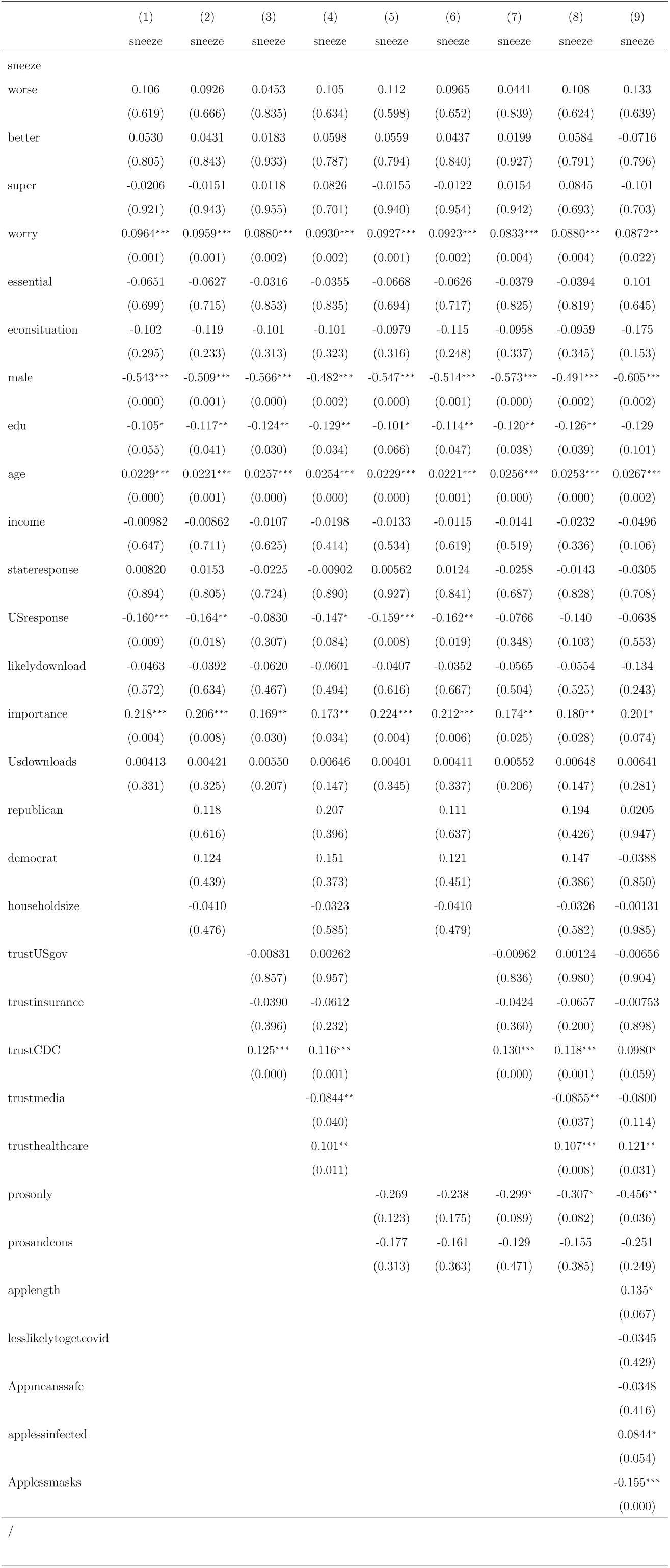
Determinants of large gatherings: combined controls, robust standard errors

**Table 9:**
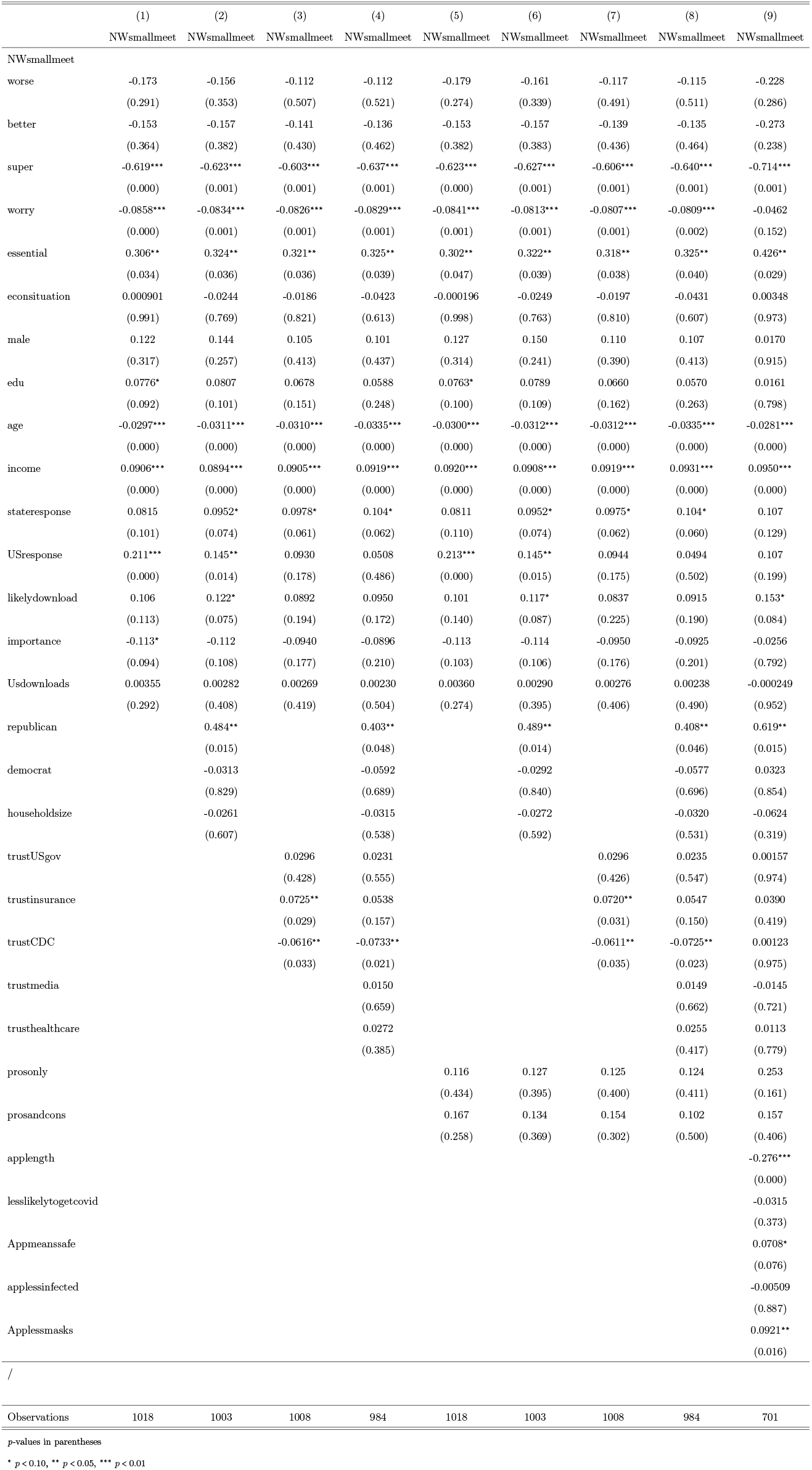
Determinants of small gatherings: combined controls, robust standard errors

**Table 10:**
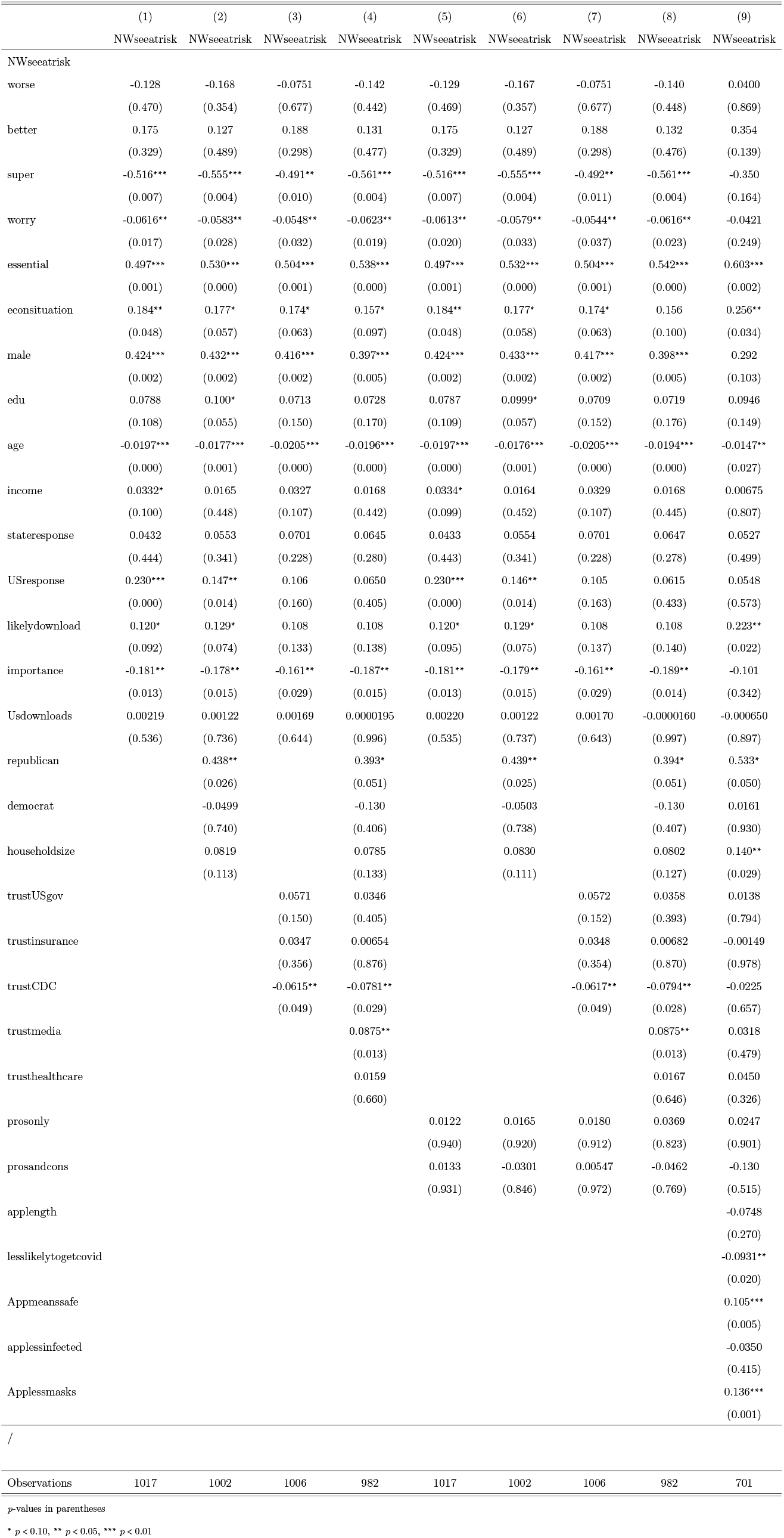
Determinants of seeing people at risk: combined controls, robust standard errors

**Table 11:**
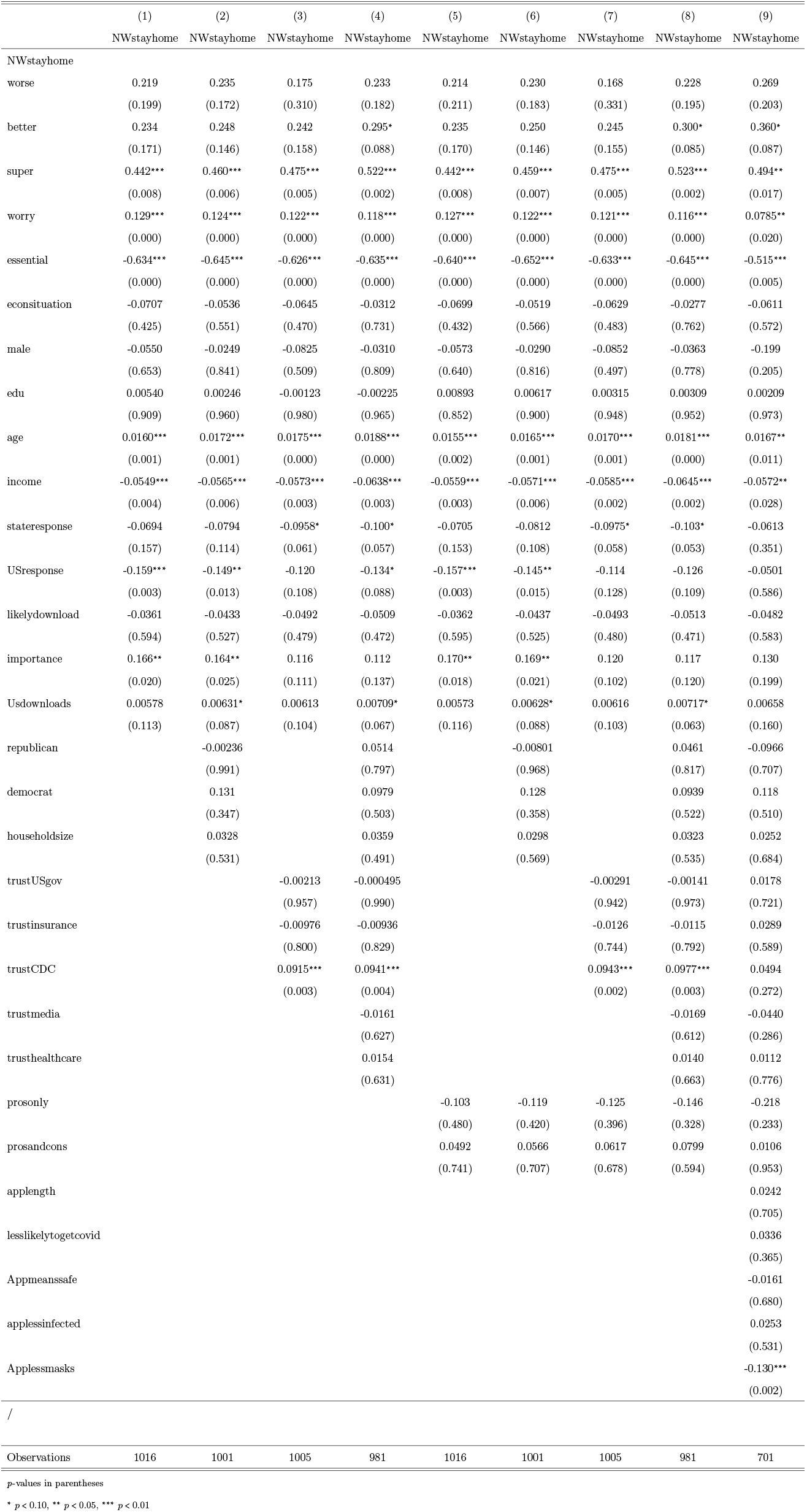
Determinants of staying home: combined controls, robust standard errors

**Table 12:** Determinants of wearing a mask: combined controls, robust standard errors

|  | (1) | (2) | (3) | (4) | (5) | (6) | (7) | (8) | (9) |
| --- | --- | --- | --- | --- | --- | --- | --- | --- | --- |
|  | NWmask | NWmask | NWmask | NWmask | NWmask | NWmask | NWmask | NWmask | NWmask |
| NWmask |  |  |  |  |  |  |  |  |  |
| worse | 0.293<br>(0.162) | 0.256<br>(0.224) | 0.185<br>(0.390) | 0.197<br>(0.362) | 0.284<br>(0.177) | 0.246<br>(0.244) | 0.172<br>(0.424) | 0.181<br>(0.401) | 0.491*<br>(0.064) |
| better | 0.341<br>(0.100) | 0.363*<br>(0.084) | 0.321<br>(0.127) | 0.390*<br>(0.070) | 0.340<br>(0.102) | 0.364*<br>(0.084) | 0.324<br>(0.124) | 0.398*<br>(0.065) | 0.509*<br>(0.078) |
| super | 0.321<br>(0.128) | 0.362*<br>(0.091) | 0.333<br>(0.117) | 0.408*<br>(0.058) | 0.313<br>(0.141) | 0.355<br>(0.100) | 0.328<br>(0.126) | 0.404*<br>(0.062) | 0.702**<br>(0.012) |
| worry | 0.219***<br>(0.000) | 0.214***<br>(0.000) | 0.209***<br>(0.000) | 0.208***<br>(0.000) | 0.225***<br>(0.000) | 0.221***<br>(0.000) | 0.215***<br>(0.000) | 0.215***<br>(0.000) | 0.156***<br>(0.000) |
| essential | -0.338**<br>(0.045) | -0.319*<br>(0.062) | -0.324*<br>(0.054) | -0.298*<br>(0.078) | -0.342**<br>(0.043) | -0.323*<br>(0.058) | -0.327*<br>(0.052) | -0.302*<br>(0.075) | -0.181<br>(0.440) |
| econsituation | -0.00222<br>(0.983) | -0.0141<br>(0.893) | 0.0193<br>(0.856) | 0.0176<br>(0.871) | -0.0133<br>(0.899) | -0.0272<br>(0.799) | 0.00887<br>(0.934) | 0.00501<br>(0.963) | -0.121<br>(0.365) |
| male | -0.500***<br>(0.002) | -0.484***<br>(0.003) | -0.529***<br>(0.001) | -0.464***<br>(0.006) | -0.500***<br>(0.002) | -0.480***<br>(0.003) | -0.528***<br>(0.001) | -0.459***<br>(0.007) | -0.397*<br>(0.079) |
| edu | -0.0733<br>(0.233) | -0.0889<br>(0.164) | -0.0772<br>(0.214) | -0.0945<br>(0.147) | -0.0761<br>(0.218) | -0.0937<br>(0.145) | -0.0779<br>(0.213) | -0.0969<br>(0.140) | -0.153*<br>(0.081) |
| age | 0.00474<br>(0.438) | 0.00419<br>(0.509) | 0.00789<br>(0.216) | 0.00642<br>(0.333) | 0.00428<br>(0.488) | 0.00370<br>(0.564) | 0.00716<br>(0.265) | 0.00566<br>(0.397) | 0.00149<br>(0.877) |
| income | 0.00414<br>(0.857) | 0.00647<br>(0.787) | 0.00743<br>(0.751) | 0.00182<br>(0.942) | 0.00916<br>(0.697) | 0.0120<br>(0.623) | 0.0122<br>(0.612) | 0.00740<br>(0.770) | -0.00245<br>(0.938) |
| stateresponse | -0.0154<br>(0.807) | -0.0174<br>(0.785) | -0.0632<br>(0.329) | -0.0600<br>(0.363) | -0.00639<br>(0.920) | -0.00659<br>(0.918) | -0.0569<br>(0.381) | -0.0510<br>(0.443) | -0.0453<br>(0.599) |
| USresponse | -0.240***<br>(0.000) | -0.193**<br>(0.011) | -0.0666<br>(0.455) | -0.0587<br>(0.523) | -0.244***<br>(0.000) | -0.198***<br>(0.010) | -0.0648<br>(0.476) | -0.0598<br>(0.525) | 0.0440<br>(0.725) |
| likelydownload | -0.00478<br>(0.955) | -0.00974<br>(0.910) | -0.0153<br>(0.863) | -0.0161<br>(0.861) | -0.0140<br>(0.871) | -0.0176<br>(0.840) | -0.0262<br>(0.773) | -0.0258<br>(0.783) | -0.0826<br>(0.489) |
| importance | 0.280***<br>(0.001) | 0.271***<br>(0.001) | 0.212**<br>(0.014) | 0.214**<br>(0.016) | 0.273***<br>(0.001) | 0.263***<br>(0.002) | 0.207**<br>(0.016) | 0.208**<br>(0.019) | 0.230*<br>(0.057) |
| Usdownloads | -0.00200<br>(0.653) | -0.00241<br>(0.598) | 0.000192<br>(0.967) | -0.000236<br>(0.961) | -0.00185<br>(0.678) | -0.00235<br>(0.609) | 0.000402<br>(0.931) | -0.000114<br>(0.981) | 0.00260<br>(0.681) |
| republican |  | 0.113<br>(0.635) |  | 0.255<br>(0.300) |  | 0.137<br>(0.567) |  | 0.285<br>(0.250) | 0.125<br>(0.708) |
| democrat |  | 0.413**<br>(0.017) |  | 0.373**<br>(0.041) |  | 0.435**<br>(0.012) |  | 0.394**<br>(0.032) | 0.322<br>(0.153) |
| householdsize |  | -0.00999<br>(0.875) |  | -0.00730<br>(0.907) |  | -0.0128<br>(0.840) |  | -0.0116<br>(0.854) | 0.0158<br>(0.842) |
| trustUSgov |  |  | -0.0560<br>(0.258) | -0.0620<br>(0.228) |  |  | -0.0581<br>(0.243) | -0.0650<br>(0.210) | -0.0946<br>(0.194) |
| trustinsurance |  |  | -0.0692<br>(0.131) | -0.100**<br>(0.037) |  |  | -0.0703<br>(0.128) | -0.0983**<br>(0.042) | 0.00430<br>(0.948) |
| trustCDC |  |  | 0.165***<br>(0.000) | 0.141***<br>(0.000) |  |  | 0.167***<br>(0.000) | 0.146***<br>(0.000) | 0.0899*<br>(0.092) |
| trustmedia |  |  |  | -0.0399<br>(0.348) |  |  |  | -0.0400<br>(0.354) | -0.0402<br>(0.499) |
| trusthealthcare |  |  |  | 0.0989**<br>(0.019) |  |  |  | 0.0949**<br>(0.025) | 0.114**<br>(0.045) |
| prosonly |  |  |  |  | 0.333*<br>(0.075) | 0.362*<br>(0.057) | 0.281<br>(0.138) | 0.320*<br>(0.098) | -0.126<br>(0.609) |
| prosandcons |  |  |  |  | 0.325*<br>(0.077) | 0.341*<br>(0.066) | 0.385**<br>(0.039) | 0.388**<br>(0.039) | 0.377<br>(0.126) |
| applength |  |  |  |  |  |  |  |  | 0.139*<br>(0.090) |
| lesslikelytogetcovid |  |  |  |  |  |  |  |  | 0.0385<br>(0.427) |
| Appmeanssafe |  |  |  |  |  |  |  |  | 0.0573<br>(0.291) |
| applessinfected |  |  |  |  |  |  |  |  | 0.0381<br>(0.464) |
| Applessmasks |  |  |  |  |  |  |  |  | -0.313***<br>(0.000) |

Overall, these results point very strongly in the direction that behavioral feedback messages can be effectively used to induce prudent behavior among app users. In particular, informing the people who are moving the most or are having the most contacts that they could be superspreaders can induce them to stay home more often, to attend less large and small gatherings and to see less people at risk. To the best of our knowledge, no app gives this information, therefore our findings provide a concrete suggestion on how to improve the effectiveness of CTAs in containing the spread of COVID-19.

Comparative messages (Good and Bad behavior) might also be helpful, as the people who receive a Good behavior message keep the app for longer (Table 13). This effect is relatively weak and not always significant, but including comparative messages should be virtually cost free, hence it might be worthwhile even if the benefit is small. Moreover, app users might respond to this type of feedback by improving their behavior in order to be recognized as being better than average.

**Table 13:** Determinants of length of keeping the app: combined controls, robust standard errors

|  | (1) | (2) | (3) | (4) | (5) | (6) | (7) | (8) | (9) |
| --- | --- | --- | --- | --- | --- | --- | --- | --- | --- |
|  | applength | applength | applength | applength | applength | applength | applength | applength | applength |
| applength |  |  |  |  |  |  |  |  |  |
| prosonly | -0.0640<br>(0.710) | -0.0320<br>(0.853) | -0.0656<br>(0.707) | -0.0236<br>(0.894) | -0.0640<br>(0.710) | -0.0320<br>(0.853) | -0.0656<br>(0.707) | -0.0236<br>(0.894) | -0.0422<br>(0.816) |
| prosandcons | -0.180<br>(0.315) | -0.173<br>(0.336) | -0.182<br>(0.316) | -0.148<br>(0.424) | -0.180<br>(0.315) | -0.173<br>(0.336) | -0.182<br>(0.316) | -0.148<br>(0.424) | -0.145<br>(0.441) |
| worse | 0.210<br>(0.314) | 0.235<br>(0.263) | 0.208<br>(0.341) | 0.247<br>(0.263) | 0.210<br>(0.314) | 0.235<br>(0.263) | 0.208<br>(0.341) | 0.247<br>(0.263) | 0.150<br>(0.496) |
| better | 0.371*<br>(0.059) | 0.401**<br>(0.044) | 0.359*<br>(0.072) | 0.400*<br>(0.052) | 0.371*<br>(0.059) | 0.401**<br>(0.044) | 0.359*<br>(0.072) | 0.400*<br>(0.052) | 0.262<br>(0.207) |
| super | 0.292<br>(0.131) | 0.321*<br>(0.099) | 0.263<br>(0.177) | 0.295<br>(0.136) | 0.292<br>(0.131) | 0.321*<br>(0.099) | 0.263<br>(0.177) | 0.295<br>(0.136) | 0.140<br>(0.503) |
| worry | 0.0416<br>(0.188) | 0.0419<br>(0.178) | 0.0332<br>(0.299) | 0.0321<br>(0.315) | 0.0416<br>(0.188) | 0.0419<br>(0.178) | 0.0332<br>(0.299) | 0.0321<br>(0.315) | 0.0280<br>(0.395) |
| essential | -0.0288<br>(0.862) | -0.0270<br>(0.871) | -0.00934<br>(0.956) | -0.00874<br>(0.960) | -0.0288<br>(0.862) | -0.0270<br>(0.871) | -0.00934<br>(0.956) | -0.00874<br>(0.960) | 0.0752<br>(0.678) |
| econsituation | 0.137<br>(0.181) | 0.164<br>(0.112) | 0.120<br>(0.250) | 0.143<br>(0.180) | 0.137<br>(0.181) | 0.164<br>(0.112) | 0.120<br>(0.250) | 0.143<br>(0.180) | 0.163<br>(0.126) |
| male | -0.143<br>(0.321) | -0.104<br>(0.469) | -0.134<br>(0.359) | -0.0642<br>(0.662) | -0.143<br>(0.321) | -0.104<br>(0.469) | -0.134<br>(0.359) | -0.0642<br>(0.662) | -0.0641<br>(0.677) |
| edu | 0.0220<br>(0.678) | 0.0440<br>(0.428) | 0.0201<br>(0.714) | 0.0535<br>(0.357) | 0.0220<br>(0.678) | 0.0440<br>(0.428) | 0.0201<br>(0.714) | 0.0535<br>(0.357) | 0.0827<br>(0.165) |
| age | -0.00333<br>(0.548) | -0.000486<br>(0.932) | -0.00534<br>(0.339) | -0.000303<br>(0.958) | -0.00333<br>(0.548) | -0.000486<br>(0.932) | -0.00534<br>(0.339) | -0.000303<br>(0.958) | -0.00219<br>(0.726) |
| income | -0.0200<br>(0.365) | -0.0347<br>(0.129) | -0.0167<br>(0.458) | -0.0328<br>(0.168) | -0.0200<br>(0.365) | -0.0347<br>(0.129) | -0.0167<br>(0.458) | -0.0328<br>(0.168) | -0.0270<br>(0.260) |
| stateresponse | -0.0342<br>(0.516) | -0.0415<br>(0.435) | -0.0291<br>(0.588) | -0.0324<br>(0.555) | -0.0342<br>(0.516) | -0.0415<br>(0.435) | -0.0291<br>(0.588) | -0.0324<br>(0.555) | -0.0539<br>(0.335) |
| USresponse | -0.0467<br>(0.534) | -0.131<br>(0.126) | 0.0156<br>(0.870) | -0.0793<br>(0.441) | -0.0467<br>(0.534) | -0.131<br>(0.126) | 0.0156<br>(0.870) | -0.0793<br>(0.441) | -0.0614<br>(0.543) |
| likelydownload | 0.296***<br>(0.001) | 0.316***<br>(0.000) | 0.297***<br>(0.001) | 0.317***<br>(0.000) | 0.296***<br>(0.001) | 0.316***<br>(0.000) | 0.297***<br>(0.001) | 0.317***<br>(0.000) | 0.318***<br>(0.001) |
| importance | 0.293***<br>(0.001) | 0.276***<br>(0.003) | 0.301***<br>(0.001) | 0.296***<br>(0.002) | 0.293***<br>(0.001) | 0.276***<br>(0.003) | 0.301***<br>(0.001) | 0.296***<br>(0.002) | 0.238**<br>(0.013) |
| Usdownloads | 0.00666<br>(0.127) | 0.00707<br>(0.105) | 0.00709<br>(0.118) | 0.00830*<br>(0.075) | 0.00666<br>(0.127) | 0.00707<br>(0.105) | 0.00709<br>(0.118) | 0.00830*<br>(0.075) | 0.0102**<br>(0.037) |
| republican |  | 0.528*<br>(0.061) |  | 0.599**<br>(0.038) |  | 0.528*<br>(0.061) |  | 0.599**<br>(0.038) | 0.591**<br>(0.046) |
| democrat |  | 0.0928<br>(0.544) |  | 0.146<br>(0.377) |  | 0.0928<br>(0.544) |  | 0.146<br>(0.377) | 0.0856<br>(0.613) |
| householdsize |  | 0.0670<br>(0.221) |  | 0.0675<br>(0.226) |  | 0.0670<br>(0.221) |  | 0.0675<br>(0.226) | 0.0971*<br>(0.099) |
| trustUSgov |  |  | -0.0653<br>(0.158) | -0.0642<br>(0.167) |  |  | -0.0653<br>(0.158) | -0.0642<br>(0.167) | -0.0451<br>(0.337) |
| trustinsurance |  |  | 0.0201<br>(0.600) | 0.0302<br>(0.479) |  |  | 0.0201<br>(0.600) | 0.0302<br>(0.479) | 0.0346<br>(0.441) |
| trustCDC |  |  | 0.00410<br>(0.911) | 0.0273<br>(0.488) |  |  | 0.00410<br>(0.911) | 0.0273<br>(0.488) | -0.00547<br>(0.894) |
| trustmedia |  |  |  | -0.0412<br>(0.263) |  |  |  | -0.0412<br>(0.263) | -0.0517<br>(0.175) |
| trusthealthcare |  |  |  | -0.0120<br>(0.729) |  |  |  | -0.0120<br>(0.729) | -0.00819<br>(0.823) |
| lesslikelytogetcovid |  |  |  |  |  |  |  |  | 0.0368<br>(0.261) |
| Appmeanssafe |  |  |  |  |  |  |  |  | -0.0770*<br>(0.066) |
| applessinfected |  |  |  |  |  |  |  |  | 0.164***<br>(0.000) |
| Applessmasks |  |  |  |  |  |  |  |  | -0.111**<br>(0.025) |
| / |  |  |  |  |  |  |  |  |  |
|  | (0.435) | (0.891) | heightObservations | 736 | 728 | 729 | 716 | 736 | 728 |
| 729 | 716 | 701 |  |  |  |  |  |  |  |

p-values in parentheses
\* $p < 0.10$ , \*\* $p < 0.05$ , \*\*\* $p < 0.01$

In line with the rest of our results we find that the effect of in-app notification depends on the characteristics of the respondents. For example, males that are in the Good behavior group will attend less small gatherings. Similarly, males that receive the Good and the Bad behavior treatment are less likely to see people at risk than males in the control group. Last, respondents in the Superspreaders group with a Professional or Master’s degree use mask much more.

However, in-app notifications sometimes triggered counterproductive responses. For instance, Republicans that see the Bad behavior treatment will see more people at risk. This result underscores the importance of tailoring in-app notifications to the characteristics of the app user.

## 6. COVID-19 Prevention Optimism

We now turn to analyze the results on the questions related with COVID-19 prevention optimism based on the CTA (Tables 1 and 14–17).

**Table 14:**
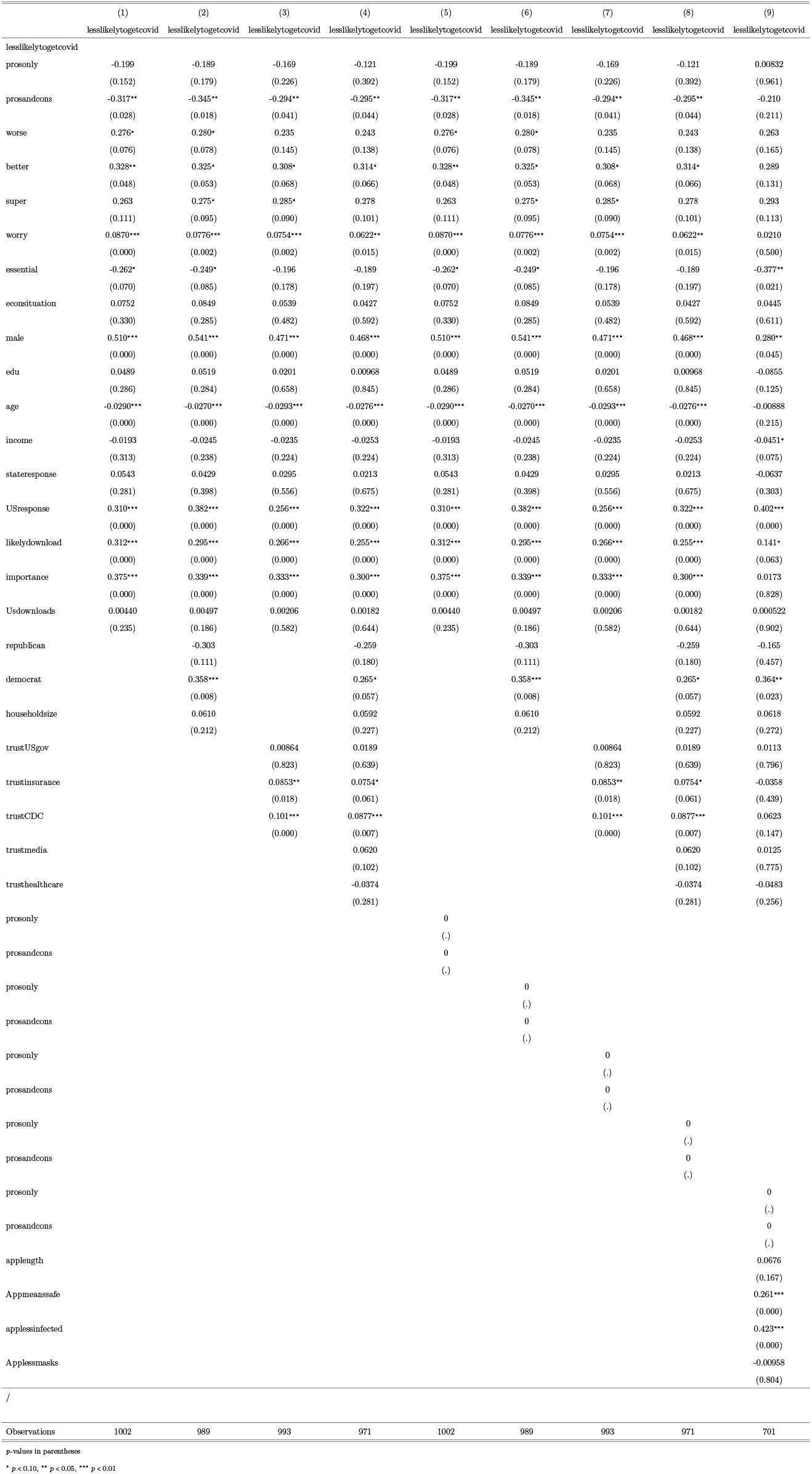
Determinants of saying you’re less likely to get covid: combined controls, robust standard errors

**Table 15:** Determinants of saying app makes us safer: combined controls, robust standard errors

|  | (1) | (2) | (3) | (4) | (5) | (6) | (7) | (8) | (9) |
| --- | --- | --- | --- | --- | --- | --- | --- | --- | --- |
|  | Appmeanssafe | Appmeanssafe | Appmeanssafe | Appmeanssafe | Appmeanssafe | Appmeanssafe | Appmeanssafe | Appmeanssafe | Appmeanssafe |
| Appmeanssafe |  |  |  |  |  |  |  |  |  |
| prosonly | -0.0542<br>(0.713) | -0.0739<br>(0.618) | -0.00217<br>(0.988) | -0.00975<br>(0.947) | -0.0542<br>(0.713) | -0.0739<br>(0.618) | -0.00217<br>(0.988) | -0.00975<br>(0.947) | 0.0824<br>(0.632) |
| prosandcons | -0.250*<br>(0.093) | -0.296*<br>(0.051) | -0.276*<br>(0.067) | -0.332**<br>(0.029) | -0.250*<br>(0.093) | -0.296*<br>(0.051) | -0.276*<br>(0.067) | -0.332**<br>(0.029) | -0.0939<br>(0.614) |
| worse | -0.373**<br>(0.020) | -0.354**<br>(0.032) | -0.318*<br>(0.053) | -0.304*<br>(0.071) | -0.373**<br>(0.020) | -0.354**<br>(0.032) | -0.318*<br>(0.053) | -0.304*<br>(0.071) | -0.622***<br>(0.002) |
| better | -0.233<br>(0.180) | -0.241<br>(0.171) | -0.215<br>(0.217) | -0.223<br>(0.203) | -0.233<br>(0.180) | -0.241<br>(0.171) | -0.215<br>(0.217) | -0.223<br>(0.203) | -0.529**<br>(0.013) |
| super | -0.355**<br>(0.032) | -0.337**<br>(0.044) | -0.286*<br>(0.085) | -0.311*<br>(0.065) | -0.355**<br>(0.032) | -0.337**<br>(0.044) | -0.286*<br>(0.085) | -0.311*<br>(0.065) | -0.554***<br>(0.004) |
| worry | 0.00783<br>(0.749) | 0.00306<br>(0.902) | 0.0108<br>(0.666) | -0.00194<br>(0.940) | 0.00783<br>(0.749) | 0.00306<br>(0.902) | 0.0108<br>(0.666) | -0.00194<br>(0.940) | -0.0524<br>(0.114) |
| essential | 0.318**<br>(0.029) | 0.324**<br>(0.028) | 0.367**<br>(0.013) | 0.378**<br>(0.014) | 0.318**<br>(0.029) | 0.324**<br>(0.028) | 0.367**<br>(0.013) | 0.378**<br>(0.014) | 0.460**<br>(0.010) |
| econsituation | 0.0908<br>(0.255) | 0.0933<br>(0.244) | 0.0478<br>(0.541) | 0.0310<br>(0.693) | 0.0908<br>(0.255) | 0.0933<br>(0.244) | 0.0478<br>(0.541) | 0.0310<br>(0.693) | -0.0535<br>(0.539) |
| male | 0.396***<br>(0.001) | 0.412***<br>(0.001) | 0.285**<br>(0.025) | 0.229*<br>(0.079) | 0.396***<br>(0.001) | 0.412***<br>(0.001) | 0.285**<br>(0.025) | 0.229*<br>(0.079) | -0.0419<br>(0.779) |
| edu | 0.107**<br>(0.026) | 0.134***<br>(0.009) | 0.0875*<br>(0.077) | 0.0914*<br>(0.085) | 0.107**<br>(0.026) | 0.134***<br>(0.009) | 0.0875*<br>(0.077) | 0.0914*<br>(0.085) | 0.0443<br>(0.480) |
| age | -0.0369***<br>(0.000) | -0.0353***<br>(0.000) | -0.0392***<br>(0.000) | -0.0398***<br>(0.000) | -0.0369***<br>(0.000) | -0.0353***<br>(0.000) | -0.0392***<br>(0.000) | -0.0398***<br>(0.000) | -0.0265***<br>(0.000) |
| income | 0.0494**<br>(0.011) | 0.0376*<br>(0.076) | 0.0394**<br>(0.048) | 0.0308<br>(0.155) | 0.0494**<br>(0.011) | 0.0376*<br>(0.076) | 0.0394**<br>(0.048) | 0.0308<br>(0.155) | 0.0254<br>(0.315) |
| stateresponse | 0.102**<br>(0.046) | 0.0965*<br>(0.063) | 0.0890*<br>(0.084) | 0.0690<br>(0.184) | 0.102**<br>(0.046) | 0.0965*<br>(0.063) | 0.0890*<br>(0.084) | 0.0690<br>(0.184) | 0.0470<br>(0.425) |
| USResponse | 0.364***<br>(0.000) | 0.374***<br>(0.000) | 0.115<br>(0.116) | 0.165**<br>(0.046) | 0.364***<br>(0.000) | 0.374***<br>(0.000) | 0.115<br>(0.116) | 0.165**<br>(0.046) | 0.000132<br>(0.999) |
| likelydownload | 0.257***<br>(0.000) | 0.243***<br>(0.001) | 0.206***<br>(0.004) | 0.180**<br>(0.013) | 0.257***<br>(0.000) | 0.243***<br>(0.001) | 0.206***<br>(0.004) | 0.180**<br>(0.013) | 0.0522<br>(0.572) |
| importance | 0.145**<br>(0.035) | 0.136*<br>(0.056) | 0.126*<br>(0.077) | 0.103<br>(0.172) | 0.145**<br>(0.035) | 0.136*<br>(0.056) | 0.126*<br>(0.077) | 0.103<br>(0.172) | -0.115<br>(0.236) |
| Usdownloads | 0.0113***<br>(0.001) | 0.0113***<br>(0.001) | 0.00789**<br>(0.022) | 0.00640*<br>(0.075) | 0.0113***<br>(0.001) | 0.0113***<br>(0.001) | 0.00789**<br>(0.022) | 0.00640*<br>(0.075) | 0.00613<br>(0.141) |
| republican |  | -0.168<br>(0.405) |  | -0.227<br>(0.267) |  | -0.168<br>(0.405) |  | -0.227<br>(0.267) | -0.275<br>(0.283) |
| democrat |  | 0.0312<br>(0.823) |  | -0.107<br>(0.468) |  | 0.0312<br>(0.823) |  | -0.107<br>(0.468) | -0.207<br>(0.212) |
| householdsize |  | 0.107**<br>(0.039) |  | 0.102*<br>(0.053) |  | 0.107**<br>(0.039) |  | 0.102*<br>(0.053) | 0.113*<br>(0.078) |
| trustUSgov |  |  | 0.0899**<br>(0.018) | 0.0852**<br>(0.029) |  |  | 0.0899**<br>(0.018) | 0.0852**<br>(0.029) | 0.0752<br>(0.106) |
| trustinsurance |  |  | 0.166***<br>(0.000) | 0.123***<br>(0.002) |  |  | 0.166***<br>(0.000) | 0.123***<br>(0.002) | 0.0321<br>(0.488) |
| trustCDC |  |  | -0.0325<br>(0.259) | -0.0682**<br>(0.031) |  |  | -0.0325<br>(0.259) | -0.0682**<br>(0.031) | -0.0696<br>(0.117) |
| trustmedia |  |  |  | 0.119***<br>(0.001) |  |  |  | 0.119***<br>(0.001) | 0.0647<br>(0.115) |
| trusthealthcare |  |  |  | 0.0162<br>(0.614) |  |  |  | 0.0162<br>(0.614) | 0.0600<br>(0.126) |
| prosonly |  |  |  |  | 0<br>(.) |  |  |  |  |
| prosandcons |  |  |  |  | 0<br>(.) |  |  |  |  |
| applength |  |  |  |  |  |  |  |  | -0.125**<br>(0.035) |
| lesslikelytogetcovid |  |  |  |  |  |  |  |  | 0.269***<br>(0.000) |
| applessinfected |  |  |  |  |  |  |  |  | 0.0682*<br>(0.062) |
| Applessmasks |  |  |  |  |  |  |  |  | 0.404***<br>(0.000) |
| / |  |  |  |  |  |  |  |  |  |
| Observations | 1004 | 991 | 993 | 971 | 1004 | 991 | 993 | 971 | 701 |

p-values in parentheses
\* $p < 0.10$ , \*\* $p < 0.05$ , \*\*\* $p < 0.01$

**Table 16:**
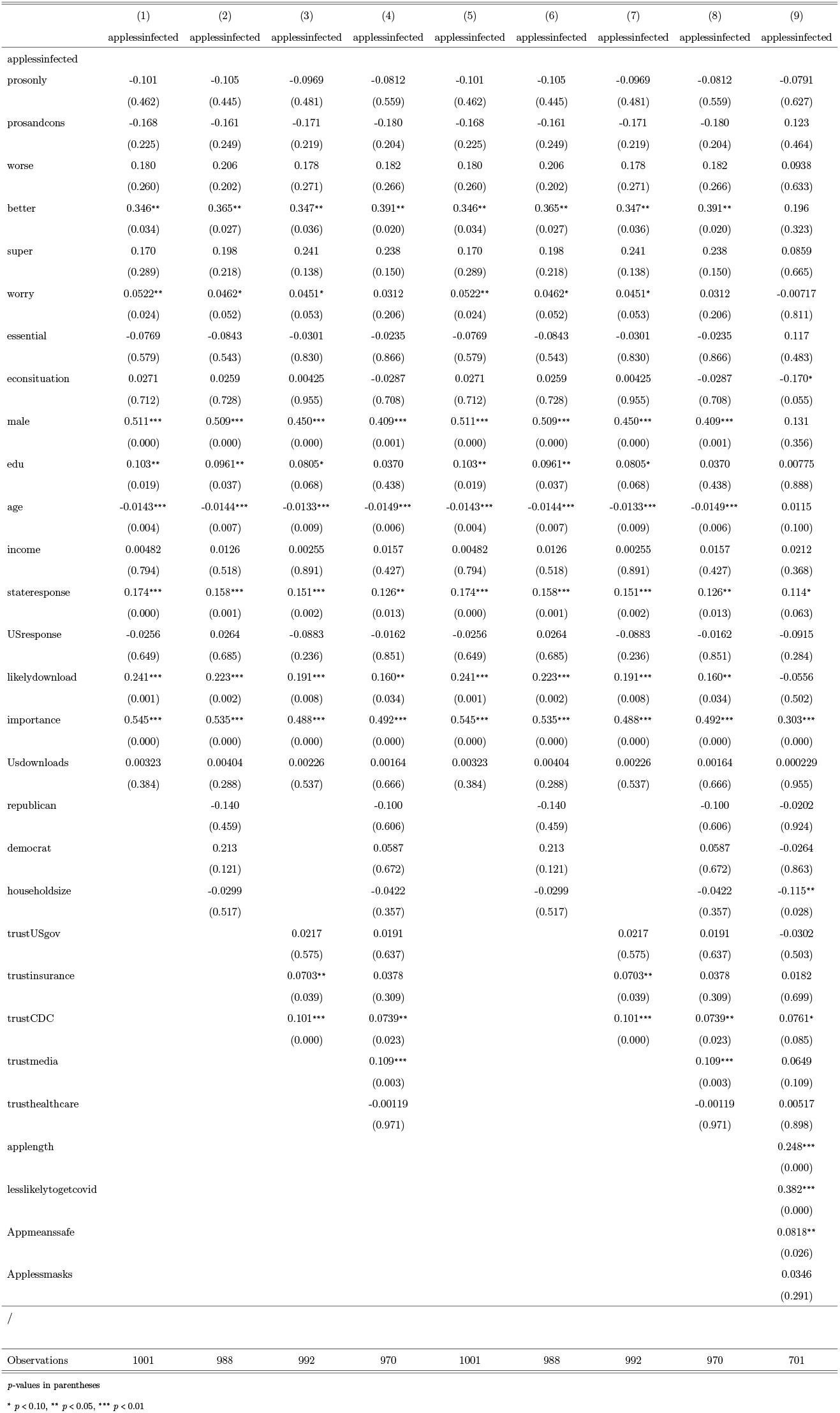
Determinants of saying app means less infected: combined controls, robust standard errors

**Table 17:**
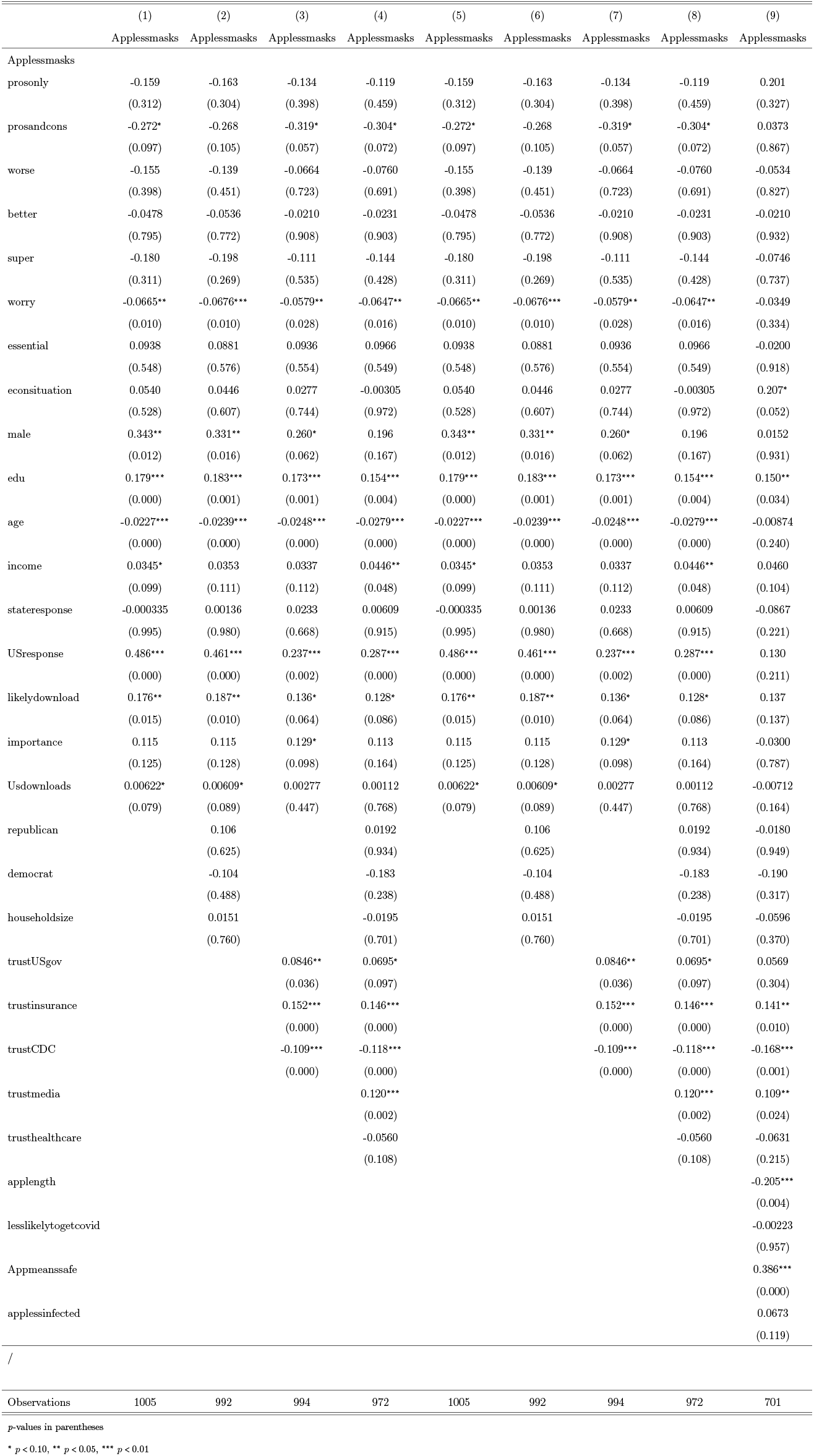
Determinants of saying app means masks are less important: combined controls, robust standard errors

These results offer some support to the idea that CTAs can cause prevention optimism for respondents that are included in the Good Behavior treatment. Vice versa, the respondents that were included in the Pros and Cons, Bad Behavior and Superspreaders treatment agree less than the Control group with some of the statements that capture prevention optimism. Consequently, a communication that is transparent about the cons of the app and in-app notifications signaling dangerous behavior create prevention *pessimism*, which could result in more prudent behavior. This confirms that in-app notifications can be effectively used to induce prudent behavior among app users. Additionally, we find that the treatments impact essential workers differently. More specifically, essential workers that are in the Bad Behavior and in the Superspreader groups agree less that the app reduces the amount of infected people and the need for wearing masks.

### 6.1. Post warning

Last, we study how much people comply with the recommendations given by the CTA after it notifies them that they have been in contact with a person who tested positive (Tables 2 and 18–20).

**Table 2:** The table summarizes the effects of the various treatments on the intention to comply with the recommendations given by the app after it notifies the user of a contact with a person who tested positive to COVID-19. * significant at 10%, ** significant at 5% and *** significant at 1%.

| **Recommendation** | **Follows More** | **Follows Less** |
| --- | --- | --- |
| Wash hands frequently with soap and water | Good Behavior** | Pros, Pros and Cons** |
| Cough and sneeze directly into a tissue or in the crook of your elbow? | - | Pros** |
| Assess your temperature at least twice daily? | - | - |
| Stay home for at least 14 days since your last exposure? | - | - |
| Maintain social distance (at least six feet) from others at all time? | - | Pros** |

**Table 3:** Frequency Table for Demographic Variables: Number, Percentage and Cumulative Percentage of respondents for the following variables: Age, Education, Income, Political orientation, Gender. Column 1 presents the statistics for the control group, Column 2 for those participants seeing only the pros about the app, Column 3 only for participants who see pros and cons about the app and Column 4 for the entire sample.

| (11em)2-10 | Control Group |  |  | Pros Only Info |  |  | Pros and Cons |  |  | Total |  |  |
| --- | --- | --- | --- | --- | --- | --- | --- | --- | --- | --- | --- | --- |
|  | No. | Col | Cum | No. | Col | Cum | No. | Col | Cum | No. | Col | Cum |
|  |  | % | % |  | % | % |  | % | % |  | % | % |
| **Age** |  |  |  |  |  |  |  |  |  |  |  |  |
| 18-25 years old | 111 | 21.9 | 21.94 | 129 | 25.6 | 25.60 | 101 | 20.0 | 19.96 | 341 | 22.5 | 22.49 |
| 26-35 years old | 162 | 32.0 | 53.95 | 152 | 30.2 | 55.75 | 163 | 32.2 | 52.17 | 477 | 31.5 | 53.96 |
| 36-45 years old | 137 | 27.1 | 81.03 | 129 | 25.6 | 81.35 | 130 | 25.7 | 77.87 | 396 | 26.1 | 80.08 |
| 46-55 years old | 46 | 9.1 | 90.12 | 41 | 8.1 | 89.48 | 50 | 9.9 | 87.75 | 137 | 9.0 | 89.12 |
| 56-65 years old | 23 | 4.5 | 94.66 | 23 | 4.6 | 94.05 | 36 | 7.1 | 94.86 | 82 | 5.4 | 94.53 |
| 66-75 years old | 16 | 3.2 | 97.83 | 18 | 3.6 | 97.62 | 15 | 3.0 | 97.83 | 49 | 3.2 | 97.76 |
| >75 years old | 11 | 2.2 | 100.00 | 12 | 2.4 | 100.00 | 11 | 2.2 | 100.00 | 34 | 2.2 | 100.00 |
| **Total** | 506 | 100.0 |  | 504 | 100.0 |  | 506 | 100.0 |  | 1516 | 100.0 |  |
| **Education** |  |  |  |  |  |  |  |  |  |  |  |  |
| Less than high school degree | 2 | 0.4 | 0.40 | 4 | 0.8 | 0.80 | 2 | 0.4 | 0.40 | 8 | 0.5 | 0.54 |
| High school graduate or equivalent | 47 | 9.5 | 9.90 | 34 | 6.8 | 7.65 | 49 | 9.8 | 10.20 | 130 | 8.7 | 9.25 |
| Some college but no degree | 107 | 21.6 | 31.52 | 105 | 21.1 | 28.77 | 100 | 20.0 | 30.20 | 312 | 20.9 | 30.16 |
| Associate degree in college (2-year) | 29 | 5.9 | 37.37 | 24 | 4.8 | 33.60 | 34 | 6.8 | 37.00 | 87 | 5.8 | 35.99 |
| Bachelor's degree in college | 173 | 34.9 | 72.32 | 179 | 36.0 | 69.62 | 158 | 31.6 | 68.60 | 510 | 34.2 | 70.17 |
| Master's or Professional Degree | 120 | 24.2 | 96.57 | 134 | 27.0 | 96.58 | 137 | 27.4 | 96.00 | 391 | 26.2 | 96.38 |
| Doctoral degree | 17 | 3.4 | 100.00 | 17 | 3.4 | 100.00 | 20 | 4.0 | 100.00 | 54 | 3.6 | 100.00 |
| **Total** | 495 | 100.0 |  | 497 | 100.0 |  | 500 | 100.0 |  | 1492 | 100.0 |  |
| **Income** |  |  |  |  |  |  |  |  |  |  |  |  |
| Less than \$10,000 | 30 | 6.1 | 6.13 | 29 | 5.8 | 5.85 | 23 | 4.7 | 4.68 | 82 | 5.6 | 5.56 |
| \$10,000 - \$19,999 | 31 | 6.3 | 12.47 | 33 | 6.7 | 12.50 | 33 | 6.7 | 11.41 | 97 | 6.6 | 12.13 |
| \$20,000 - \$29,999 | 39 | 8.0 | 20.45 | 47 | 9.5 | 21.98 | 28 | 5.7 | 17.11 | 114 | 7.7 | 19.85 |
| \$30,000 - \$39,999 | 47 | 9.6 | 30.06 | 41 | 8.3 | 30.24 | 57 | 11.6 | 28.72 | 145 | 9.8 | 29.67 |
| \$40,000 - \$49,999 | 36 | 7.4 | 37.42 | 43 | 8.7 | 38.91 | 46 | 9.4 | 38.09 | 125 | 8.5 | 38.14 |
| \$50,000 - \$59,999 | 39 | 8.0 | 45.40 | 50 | 10.1 | 48.99 | 42 | 8.6 | 46.64 | 131 | 8.9 | 47.02 |
| \$60,000 - \$69,999 | 36 | 7.4 | 52.76 | 30 | 6.0 | 55.04 | 38 | 7.7 | 54.38 | 104 | 7.0 | 54.07 |
| \$70,000 - \$79,999 | 34 | 7.0 | 59.71 | 37 | 7.5 | 62.50 | 40 | 8.1 | 62.53 | 111 | 7.5 | 61.59 |
| \$80,000 - \$89,999 | 16 | 3.3 | 62.99 | 20 | 4.0 | 66.53 | 22 | 4.5 | 67.01 | 58 | 3.9 | 65.51 |
| \$80,000 - \$89,999 | 33 | 6.7 | 69.73 | 28 | 5.6 | 72.18 | 27 | 5.5 | 72.51 | 88 | 6.0 | 71.48 |
| \$90,000 - \$99,999 | 100 | 20.4 | 90.18 | 105 | 21.2 | 93.35 | 91 | 18.5 | 91.04 | 296 | 20.1 | 91.53 |
| \$150,000 or more | 48 | 9.8 | 100.00 | 33 | 6.7 | 100.00 | 44 | 9.0 | 100.00 | 125 | 8.5 | 100.00 |
| **Total** | 489 | 100.0 |  | 496 | 100.0 |  | 491 | 100.0 |  | 1476 | 100.0 |  |
| **Political Orientation** |  |  |  |  |  |  |  |  |  |  |  |  |
| Other | 28 | 5.5 | 5.53 | 32 | 6.3 | 6.35 | 26 | 5.1 | 5.14 | 86 | 5.7 | 5.67 |
| Democrat | 229 | 45.3 | 50.79 | 229 | 45.4 | 51.79 | 228 | 45.1 | 50.20 | 686 | 45.3 | 50.92 |
| Republican | 119 | 23.5 | 74.31 | 108 | 21.4 | 73.21 | 118 | 23.3 | 73.52 | 345 | 22.8 | 73.68 |

**Table 4:** Summary statistics (mean and standard deviation (SD)) for the variables considered in the regression tables. These include: Worry About Covid-19, stay at home whenever possible, attendance of small meetings, attendance of large meetings, seeing people at risk, likelihood to wear a mask whenever going out in the next week, gender, being an essential worker, education, economic situation, age, republican, democrat, and all the measures of trust: in people known by the respondent, people not known to the respondent, in the media, in insurance companies, in the healthcare system, in the US government and in the CDC. Column 1 presents the statistics for the control group, Column 2 for those participants seeing only the pros about the app, Column 3 only for participants who see pros and cons about the app and Column 4 for the entire sample.

|  | (All Sample) |  | (Pros Only) |  | (Pros and Cons) |  | (Control) |  |
| --- | --- | --- | --- | --- | --- | --- | --- | --- |
|  | mean | sd | mean | sd | mean | sd | mean | sd |
| Worry About Covid-19 | 6.48 | 2.90 | 6.33 | 2.93 | 6.52 | 2.84 | 6.60 | 2.93 |
| Stay at home | 7.97 | 2.55 | 7.93 | 2.59 | 7.90 | 2.59 | 8.09 | 2.46 |
| Attend Small Meetings | 3.44 | 3.53 | 3.41 | 3.59 | 3.72 | 3.58 | 3.19 | 3.42 |
| Attend Large Meetings | 1.66 | 2.95 | 1.62 | 2.96 | 1.82 | 3.04 | 1.54 | 2.83 |
| See People at Risk | 2.76 | 3.38 | 2.81 | 3.47 | 2.91 | 3.47 | 2.56 | 3.19 |
| Likelihood to Wear Masks Next Week | 8.99 | 2.06 | 9.15 | 1.80 | 8.89 | 2.20 | 8.92 | 2.16 |
| Male | 0.54 | 0.50 | 0.53 | 0.50 | 0.54 | 0.50 | 0.54 | 0.50 |
| Essential worker | 0.30 | 0.46 | 0.27 | 0.45 | 0.32 | 0.47 | 0.30 | 0.46 |
| Education | 4.58 | 1.41 | 4.63 | 1.38 | 4.58 | 1.44 | 4.52 | 1.41 |
| Economic situation | 2.72 | 0.90 | 2.73 | 0.89 | 2.73 | 0.89 | 2.71 | 0.91 |
| Age | 36.04 | 12.60 | 35.40 | 12.71 | 36.91 | 12.82 | 35.82 | 12.25 |
| Republican | 0.23 | 0.42 | 0.21 | 0.41 | 0.23 | 0.42 | 0.24 | 0.42 |
| Democrat | 0.45 | 0.50 | 0.45 | 0.50 | 0.45 | 0.50 | 0.45 | 0.50 |
| Trust: People known | 8.05 | 1.76 | 8.16 | 1.63 | 8.09 | 1.83 | 7.91 | 1.82 |
| Trust: People not known | 3.54 | 2.63 | 3.58 | 2.64 | 3.64 | 2.73 | 3.40 | 2.52 |
| Trust: Media | 4.14 | 2.83 | 4.14 | 2.83 | 4.23 | 2.87 | 4.04 | 2.80 |
| Trust: Insurance | 3.99 | 2.90 | 3.90 | 2.91 | 4.18 | 2.90 | 3.88 | 2.89 |
| Trust: Healthcare | 5.59 | 2.73 | 5.60 | 2.65 | 5.85 | 2.74 | 5.32 | 2.78 |
| Trust: US government | 3.41 | 3.06 | 3.36 | 3.08 | 3.52 | 3.09 | 3.34 | 3.03 |
| Trust: CDC | 6.77 | 2.51 | 6.97 | 2.36 | 6.74 | 2.60 | 6.59 | 2.55 |
| Observations | 1516 |  | 504 |  | 506 |  | 506 |  |

**Table 18:**
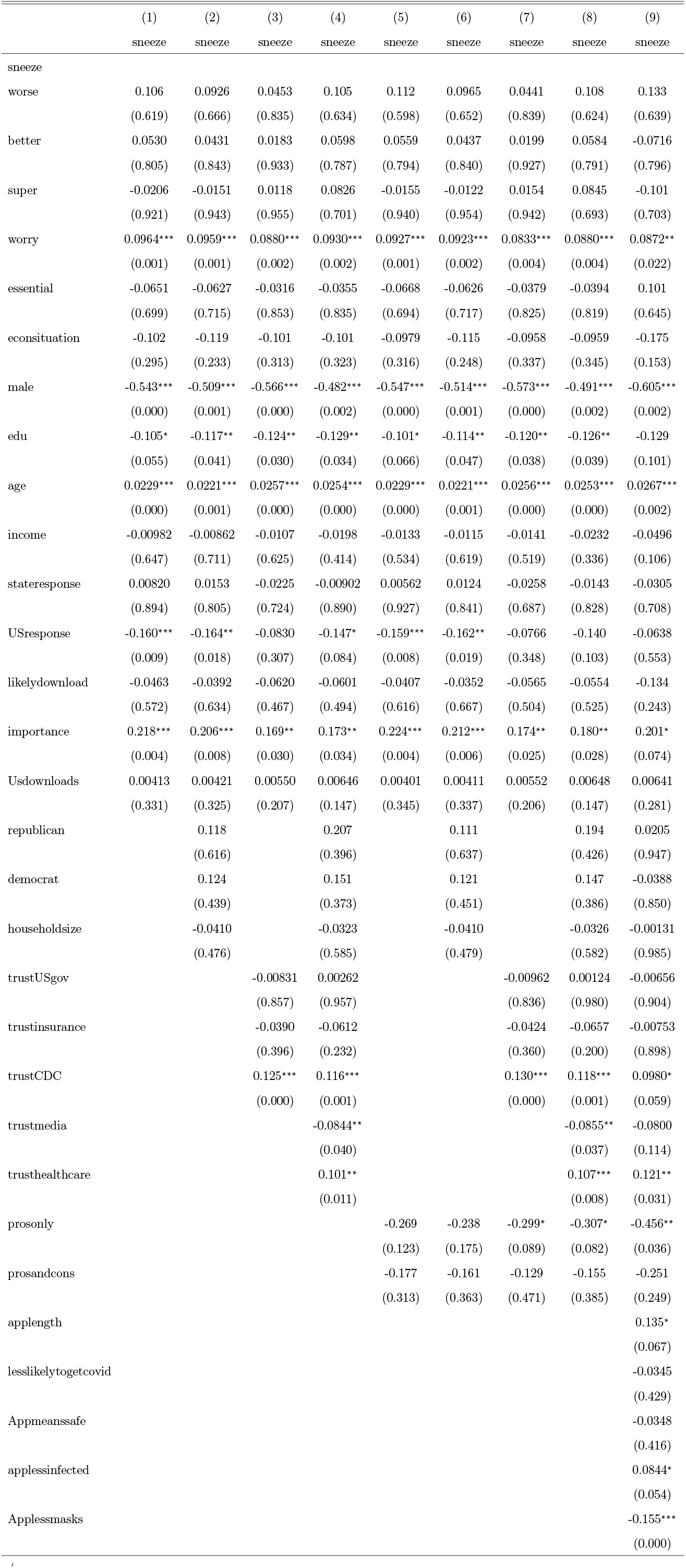
Determinants of sneezing in elbow: combined controls, robust standard errors

**Table 19:**
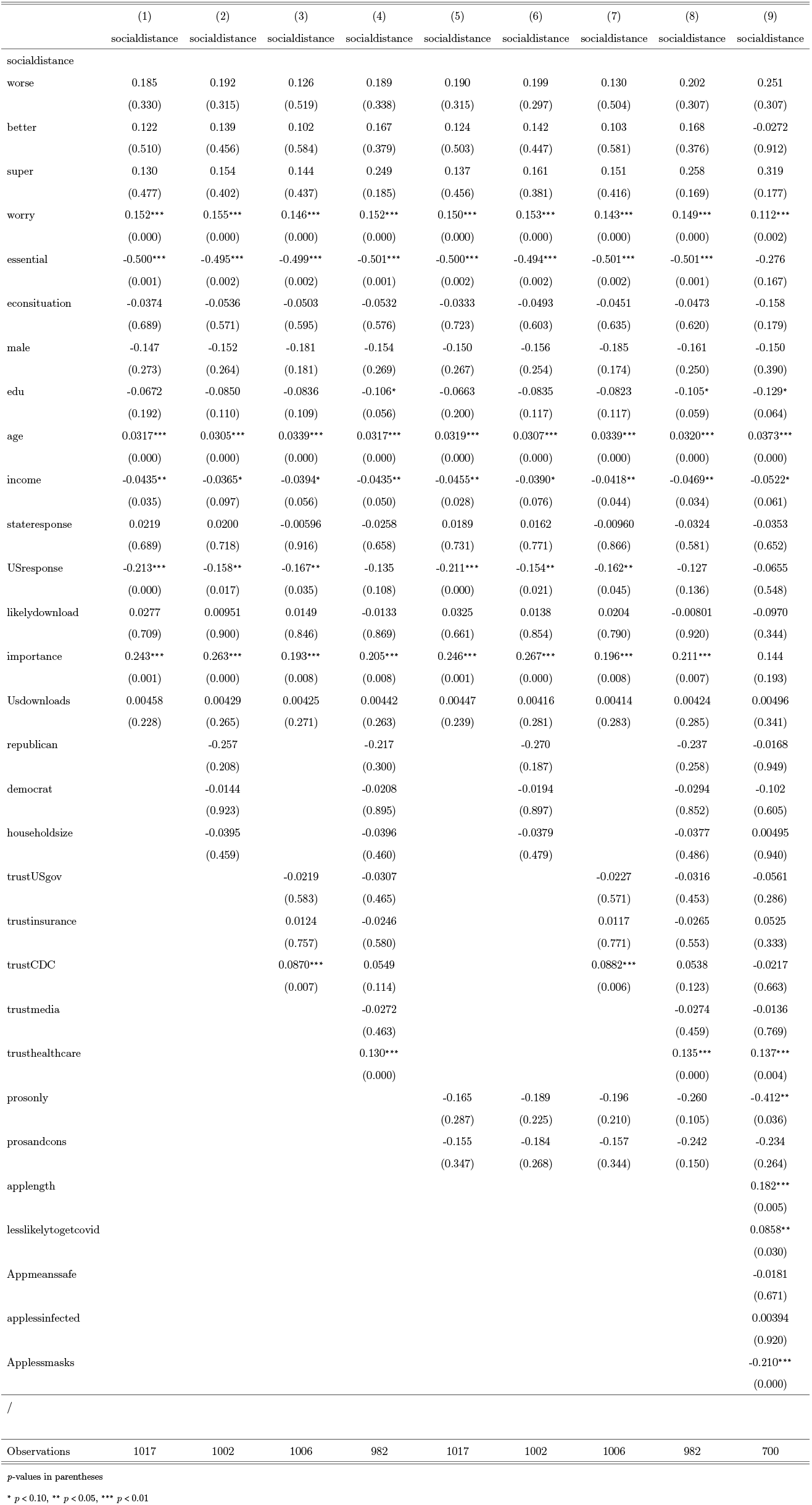
Determinants of social distancing after warning: combined controls, robust standard errors

**Table 20:** Determinants of washing hands: combined controls, robust standard errors

|  | (1) | (2) | (3) | (4) | (5) | (6) | (7) | (8) | (9) |
| --- | --- | --- | --- | --- | --- | --- | --- | --- | --- |
|  | washhands | washhands | washhands | washhands | washhands | washhands | washhands | washhands | washhands |
| washhands |  |  |  |  |  |  |  |  |  |
| worse | 0.264<br>(0.193) | 0.225<br>(0.270) | 0.220<br>(0.281) | 0.235<br>(0.253) | 0.289<br>(0.154) | 0.250<br>(0.222) | 0.242<br>(0.237) | 0.264<br>(0.201) | 0.321<br>(0.236) |
| better | 0.517**<br>(0.016) | 0.507**<br>(0.020) | 0.491**<br>(0.024) | 0.519**<br>(0.019) | 0.530**<br>(0.014) | 0.517**<br>(0.017) | 0.499**<br>(0.022) | 0.526**<br>(0.017) | 0.662**<br>(0.031) |
| super | -0.0114<br>(0.955) | 0.00158<br>(0.994) | 0.00945<br>(0.963) | 0.0712<br>(0.729) | -0.00140<br>(0.994) | 0.00970<br>(0.962) | 0.0155<br>(0.939) | 0.0756<br>(0.713) | -0.0911<br>(0.716) |
| worry | 0.130***<br>(0.000) | 0.129***<br>(0.000) | 0.123***<br>(0.000) | 0.130***<br>(0.000) | 0.127***<br>(0.000) | 0.126***<br>(0.000) | 0.119***<br>(0.000) | 0.126***<br>(0.000) | 0.0851**<br>(0.020) |
| essential | -0.335**<br>(0.048) | -0.329*<br>(0.055) | -0.309*<br>(0.071) | -0.320*<br>(0.061) | -0.327*<br>(0.056) | -0.316*<br>(0.069) | -0.306*<br>(0.075) | -0.311*<br>(0.072) | -0.373*<br>(0.093) |
| econsituation | -0.0756<br>(0.479) | -0.0813<br>(0.449) | -0.0643<br>(0.551) | -0.0497<br>(0.647) | -0.0675<br>(0.526) | -0.0742<br>(0.488) | -0.0567<br>(0.599) | -0.0432<br>(0.689) | -0.114<br>(0.382) |
| male | -0.328**<br>(0.032) | -0.333**<br>(0.030) | -0.327**<br>(0.037) | -0.277*<br>(0.080) | -0.338**<br>(0.028) | -0.344**<br>(0.026) | -0.340**<br>(0.032) | -0.290*<br>(0.070) | -0.406**<br>(0.044) |
| edu | -0.0520<br>(0.378) | -0.0537<br>(0.387) | -0.0698<br>(0.251) | -0.0562<br>(0.386) | -0.0510<br>(0.387) | -0.0519<br>(0.402) | -0.0685<br>(0.259) | -0.0543<br>(0.404) | -0.0267<br>(0.754) |
| age | 0.0152**<br>(0.019) | 0.0166**<br>(0.015) | 0.0181***<br>(0.008) | 0.0197***<br>(0.007) | 0.0157**<br>(0.016) | 0.0174**<br>(0.012) | 0.0185***<br>(0.007) | 0.0204***<br>(0.006) | 0.0217**<br>(0.035) |
| income | -0.0204<br>(0.347) | -0.0251<br>(0.287) | -0.0219<br>(0.323) | -0.0366<br>(0.135) | -0.0239<br>(0.270) | -0.0295<br>(0.212) | -0.0253<br>(0.251) | -0.0411*<br>(0.093) | -0.0441<br>(0.153) |
| stateresponse | -0.0218<br>(0.725) | -0.0203<br>(0.748) | -0.0633<br>(0.329) | -0.0501<br>(0.452) | -0.0261<br>(0.674) | -0.0245<br>(0.697) | -0.0664<br>(0.305) | -0.0549<br>(0.412) | -0.0327<br>(0.701) |
| USresponse | -0.146**<br>(0.024) | -0.132*<br>(0.073) | -0.0173<br>(0.845) | -0.0626<br>(0.502) | -0.147**<br>(0.022) | -0.134*<br>(0.068) | -0.0198<br>(0.823) | -0.0644<br>(0.489) | -0.0118<br>(0.926) |
| likelydownload | 0.114<br>(0.216) | 0.111<br>(0.233) | 0.104<br>(0.274) | 0.109<br>(0.267) | 0.126<br>(0.170) | 0.122<br>(0.191) | 0.116<br>(0.219) | 0.119<br>(0.218) | 0.0804<br>(0.538) |
| importance | 0.106<br>(0.201) | 0.0962<br>(0.255) | 0.0492<br>(0.567) | 0.0572<br>(0.528) | 0.105<br>(0.211) | 0.0964<br>(0.261) | 0.0494<br>(0.570) | 0.0592<br>(0.520) | 0.0429<br>(0.745) |
| Usdownloads | 0.00244<br>(0.559) | 0.00202<br>(0.631) | 0.00353<br>(0.407) | 0.00441<br>(0.309) | 0.00229<br>(0.582) | 0.00183<br>(0.664) | 0.00335<br>(0.433) | 0.00416<br>(0.339) | 0.00644<br>(0.286) |
| republican |  | 0.00151<br>(0.995) |  | 0.106<br>(0.655) |  | -0.00414<br>(0.986) |  | 0.0884<br>(0.712) | -0.242<br>(0.438) |
| democrat |  | 0.140<br>(0.408) |  | 0.183<br>(0.303) |  | 0.134<br>(0.428) |  | 0.177<br>(0.321) | 0.0351<br>(0.871) |
| householdsize |  | 0.0174<br>(0.775) |  | 0.0345<br>(0.583) |  | 0.0250<br>(0.685) |  | 0.0400<br>(0.528) | 0.0447<br>(0.579) |
| trustUSgov |  |  | -0.0754<br>(0.113) | -0.0670<br>(0.172) |  |  | -0.0752<br>(0.114) | -0.0666<br>(0.174) | -0.0574<br>(0.354) |
| trustinsurance |  |  | 0.0103<br>(0.814) | -0.00140<br>(0.977) |  |  | 0.0120<br>(0.787) | -0.00227<br>(0.963) | 0.0261<br>(0.686) |
| trustCDC |  |  | 0.119***<br>(0.001) | 0.113***<br>(0.003) |  |  | 0.118***<br>(0.001) | 0.109***<br>(0.005) | 0.0513<br>(0.372) |
| trustmedia |  |  |  | -0.0988**<br>(0.023) |  |  |  | -0.101**<br>(0.019) | -0.108**<br>(0.041) |
| trusthealthcare |  |  |  | 0.0848**<br>(0.021) |  |  |  | 0.0921**<br>(0.014) | 0.142***<br>(0.007) |
| prosonly |  |  |  |  | -0.312*<br>(0.085) | -0.309*<br>(0.092) | -0.333*<br>(0.069) | -0.354*<br>(0.056) | -0.481**<br>(0.032) |
| prosandcons |  |  |  |  | -0.435**<br>(0.016) | -0.466**<br>(0.010) | -0.385**<br>(0.036) | -0.447**<br>(0.016) | -0.492**<br>(0.043) |
| applength |  |  |  |  |  |  |  |  | 0.142*<br>(0.058) |
| lesslikelytogetcovid |  |  |  |  |  |  |  |  | 0.0865*<br>(0.053) |
| Appmeanssafe |  |  |  |  |  |  |  |  | -0.0387<br>(0.400) |
| applessinfected |  |  |  |  |  |  |  |  | -0.0346<br>(0.482) |
| Applessmasks |  |  |  |  |  |  |  |  | -0.237***<br>(0.000) |
| / |  |  |  |  |  |  |  |  |  |
| Observations | 1019 | 1004 | 1008 | 984 | 1019 | 1004 | 1008 | 984 | 701 |

*p*-values in parentheses\* $p < 0.10$ , \*\* $p < 0.05$ , \*\*\* $p < 0.01$

We find that our results are consistent with standard findings of the literature. For instance, male subjects reported that they would be less likely to sneeze into the crook of their elbows (*p* < 0.01) and generally speaking reported that they would take more risk.

Also in this case the impact of different messages depends on the characteristics of the app user. We find that essential workers in the Bad Behavior group are more likely to wash their hands often, to measure more frequently their temperature and to respect social distancing. The level of trust that participants declare also affects how they react to these recommendations. For instance, respondents with high trust state that they would respect more social distancing, would assess their temperature more often and would sneeze more frequently in the crook of their elbow. We find also that younger people in the Good Behavior, Bad Behavior and Superspreaders group will assess their temperatures less often and those in the Good Behavior group stay home less.

## 7. Discussion

In most countries the penetration rate of CTAs is extremely low, and hence their success has been limited. Is this the end of the story for CTAs? Not necessarily. Our results suggest a possible way forward based on the insights of behavioral economics.

To begin with, information on the CTA reduces how worried people are about the pandemic, without increasing the amount of risky activities in which they engage (Table 5). Given the devastating impact that COVID-19 is having on the mental well-being of many people [51, 52], the importance of this effect should not be underestimated. Reducing how worried people are about the pandemic is in itself a desirable goal, especially when this reduction is not accompanied by increased risk taking.

Second, we find that a more transparent communication that also emphasizes the downsides of the CTA can increase the number of downloads (Table 6). One potential explanation for this finding is that people find more credible a communication that also touches on the weaknesses of the app. To put it differently, authorities might be more credible when they guarantee that CTAs protect privacy if they have been transparent about problems like false positive and false negatives that are likely to constitute an issue [19].

However, the most important finding of our study is that providing users with useful information, such as in-app notifications on their current level of risk-taking, can significantly alter people’s behaviors. Respondents that were included in the Superspreader group were significantly less likely to attend large and small gatherings and to see people at risk, while they were significantly more likely to stay home. This suggests that CTAs can play an important role in promoting pro-social behavior during the pandemic. While also this function is best carried out when many people download the app, it does not require a minimum number of downloads. For instance, consider the case of France. As roughly 3% of the population (about 2 million people) downloaded the CTA, the number of contacts that it can identify is likely to be minimal because the likelihood that two people (infected and potentially infected) that meet both have the app is extremely small. In fact, in the first three weeks of its existence the app only notified 18 people that they had been exposed to COVID-19 [12]. This number is too small to have a large impact on how the virus spreads. However, inducing 2 million people to take less risk via tailored in-app messages could help containing the spread of COVID-19.

Consequently, we argue that CTAs should be re-framed as Behavioral Feedback Apps and their main function should be providing users with useful information on how to behave to minimize the risk of being infected with COVID-19. Contact tracing should then become an ancillary function of these BFAs, and it should be performed only for users that decide to activate it.

The BFA could indicate how crowded an area is likely to be based on information like the percentage of hotels and bed and breakfasts booked on platforms like Booking.com and AirBnB.com. Another important function that the BFA could perform is a rating system. Similarly to what apps like TripAdvisor do, the BFA might allow users to rate businesses based on how much they respect the rules on mask use, social distancing, and so on. This would leverage the wisdom of the crowd to identify the safest businesses, and might push business owners and employees to follow safety norms more closely. Online reviews have already been proposed as a tool to determine in which restaurants hygiene inspections should be carried out [50]. The wisdom of the crowd might be even more effective in this context since it is very easy to determine whether people inside a store are wearing a mask, whereas for a customer it is hard to assess the level of hygiene of a restaurant’s kitchen. Therefore, businesses’ ratings and information on which areas are safer to visit can offer precious guidance to the app users, and could help public authorities identifying the businesses that systematically violate safety norms. Moreover, receiving valuable information from the app could contribute to create a bond between the users and the app, which might increase the trust that the user has in the app.

BFAs should also have a tracing function akin to the one that CTAs currently have, and the user should be allowed to activate it at any moment. If the user chooses to do so, the app should also keep track of how much a user has moved – or how many contacts she has had – compared to similar users. This way, the app would be able to give tailored advice such as “You are taking less risks than your peers. You are protecting your community” or “You are among the users that took the most risks. You could become a superspreader”. Our results show that behavioral feedback messages of this kind can effectively induce people to engage in more prudent behavior.

## 8. Future Research

CTAs are failing in virtually every country in which they have been introduced. Scientists are attempting to find better technology that could marginally reduce the privacy risks or the number of false positives and false negatives. We argue that the reasons behind this failure lie elsewhere, and marginal technological improvements alone cannot turn the table around. CTAs touch fundamental individual rights, and their success ultimately depends on the fact that people recognize their importance and accept to use them. Consequently, the way forward is to look at behavioral sciences.

We have shown that in-app notifications based on the insights of behavioral sciences can lead to more prudent behavior. But behavioral sciences can inform many aspects of CTAs development, as even small differences in how COVID-19 related information is presented can have a big impact on how people react [53]. Consequently, we suggest that the CTAs should be abandoned in favor of Behavioral Feedback Apps. To begin with, behavioral Feedback App is a less threatening name that does not immediately evoke monitoring by the government. Moreover, it would be more appropriate to describe the functioning of the apps as we envisage them, as contact tracing, besides being entirely optional, would be only *one* of the functions performed by apps. Future studies could test whether this re-branding of the app could increase the number of downloads, and how often people using a BFA would activate a tracing function.

Additionally, it is possible to devise mechanisms of carrots and sticks to incentivize users to activate the tracing function. For example, as a carrot the governments could partner with businesses to provide incentives and discounts to customers that activated contact tracing when they visit stores. As a stick, stores and restaurants could be given the possibility to refuse entrance to customers that do not have the contact tracing function activated. This can be achieved via a Bluetooth Handshake that has already been introduced in some CTAs [19]. While this solution would push people to install the app, it is likely to be perceived very differently from a government imposition. In fact, if the adoption came from businesses it could be interpreted by many people as the sign that business owners care for the safety of their employees and customers and could contribute to create a climate of trust among people, and between people and the app. On the contrary, a similar rule introduced by the government would be seen in many countries as an unacceptable violation of freedom.

## Data Availability

All anonymized data are available from the authors upon request and will be published along with the final article

## Acknowledgments

We are very grateful to Pranjal Drall for the invaluable research assistance.

